# Using ECG Machine Learning for Detection of Cardiovascular Disease in African American Men and Women: the Jackson Heart Study

**DOI:** 10.1101/2020.07.02.20145128

**Authors:** James D. Pollard, Kazi T. Haq, Katherine J. Lutz, Nichole M. Rogovoy, Kevin A. Paternostro, Elsayed Z. Soliman, Joseph Maher, João A.C. Lima, Solomon Musani, Larisa G. Tereshchenko

## Abstract

**Background:** Almost half of African American (AA) men and women have cardiovascular disease (CVD). Detection of prevalent CVD in barbershops would facilitate secondary prevention of CVD. We sought to investigate the cross-sectional association of prevalent CVD and sex with global electrical heterogeneity (GEH) and develop a tool for CVD detection.

**Methods:** Participants from the Jackson Heart Study (JHS) with analyzable ECGs (n=3,679; age, 62±12 years; 36% men) were included. QRS, T, and spatial ventricular gradient (SVG) vectors’ magnitude and direction, and traditional metrics were measured on 12-lead ECG. Linear regression and mixed linear models with random intercept were adjusted for cardiovascular risk factors, sociodemographic and anthropometric characteristics, type of median beat, and mean RR’ intervals. Random forests, convolutional neural network, and lasso models were developed in 80%, and validated in 20% samples.

**Results:** In fully adjusted models, women had a smaller spatial QRS-T angle (−12.2(−19.4 to-5.1)°; *P*=0.001), SAI QRST (−29.8(−39.3 to −20.3) mV*ms; *P*<0.0001), and SVG elevation (−4.5(−7.5 to −1.4)°; *P*=0.004) than men, but larger SVG azimuth (+16.2(10.5-21.9)°; *P*<0.0001), with a significant random effect between families (+20.8(8.2-33.5)°; *P*=0.001). SAI QRST was larger in women with CVD as compared to CVD-free women or men (+15.1(3.8-26.4) mV*ms; *P*=0.009). Men with CVD had smaller T area [by 5.1 (95%CI 1.2-9.0) mV*ms] than CVD-free men, but there were no differences when comparing women with CVD to CVD-free women. Machine-learning detected CVD with ROC AUC 0.69-0.74; plug-in-based model included only age and QRS-T angle.

**Conclusions:** GEH varies by sex. Sex modifies an association of GEH with CVD. Automated CVD detection is feasible.

## Introduction

Almost half of African American (AA) men and women have some form of cardiovascular disease (CVD)^1^. Notable racial disparities in CVD prevalence, management, and outcomes have persisted for decades.^2^ In the United States (US), only 6% of medical school graduates are AAs, indicating a possibility of unconscious bias against AA patients. Perceived discrimination experienced by AAs is associated with mistrust of healthcare providers, negatively impacting continuity of care and adherence to treatment.^3^ In AA communities, health outreach to barbershops is common.^4^ Recent randomized controlled trial (RCT) showed that pharmacist-led treatment of hypertension in barbershops produces larger blood pressure (BP) reduction, as compared with standard BP management provided by primary care practices.^5^ Sustained effect of community-based intervention^6^ generated further ideas for pharmacist-led CVD management.^7^

Up to one-half of acute myocardial infarctions (MI) are missed or unrecognized at the time of the event, but ultimately cause heart failure (HF)^8^ or sudden cardiac death (SCD).^9^ An electrocardiogram (ECG) is one of the simplest, cheapest, and most widely available methods used to evaluate the heart. While ECG diagnosis of MI requires a physician’s interpretation, there are a growing number of automated algorithms analyzing ECG in smartphones and mobile devices. Detection of prevalent CVD in barbershops can potentially open an opportunity for secondary prevention of CVD^10^ in AA patients who have limited access to medical care. Still, it is unclear how accurately ECG can detect prevalent CVD.

While racial^11^ and sex differences^12^ in ECG characteristics have been previously described, sex differences in the association of prevalent CVD with ECG phenotype have been studied mostly in white persons. Global electrical heterogeneity (GEH)^13^ is a novel vectorcardiographic (VCG) phenotype providing additional predictive value beyond traditional ECG metrics.^14^ GEH is associated with SCD^15^, cardiovascular mortality^16^, and left ventricular dysfunction^17^ after rigorous adjustment for known cardiovascular risk factors. Sex differences in GEH have been shown in predominantly white populations.^12, 16^ However, sex differences in GEH and an association of prevalent CVD with GEH in AA men and women have not been previously studied.

To address listed above knowledge gaps, we conducted a cross-sectional study of GEH in AA participants of the Jackson Heart Study (JHS) with two goals: (1) investigate the cross-sectional associations of prevalent CVD and sex with GEH, and (2) develop and validate a tool for detection of prevalent CVD on 12-lead ECG. We hypothesized that (1) the prevalent CVD is associated with GEH after adjustment for demographic, anthropometric, socioeconomic, and traditional cardiovascular risk factors, (2) there are sex differences in GEH, and (3) sex modifies an association of prevalent CVD with GEH. We also hypothesized that automated 12-lead ECG analysis can be used to detect prevalent CVD.

## Methods

The JHS data are available through the National Heart, Lung, and Blood Institute’s Biological Specimen and Data Repository Information Coordinating Center (BioLINCC)^18^ and the National Center of Biotechnology Information’s database of Genotypes and Phenotypes (dbGaP).^19^ All study participants provided written informed consent before entering the JHS study. This study was approved by the Oregon Health & Science University (OHSU) Institutional Review Board.

### Study population

The JHS was initiated in 1998 as a prospective cohort study of CVD in AAs.^20, 21^ The JHS enrolled 5,306 participants from the Jackson, Mississippi metropolitan area from 2000-2004. Recruitment strategies included: (1) enrollment of the Atherosclerosis Risk in Communities (ARIC)^22^ study participants, (2) random and (3) volunteer recruitment pools, and (4) enrollment of secondary family members. Eligible participants were 35-84 years of age, except in a nested family cohort, which included younger participants (21-84 years of age).

In this cross-sectional study, we included JHS participants who had analyzable resting 12-lead ECG recorded as a part of the third clinical examination in 2009-2013 (Figure 1; n=3,717). We further excluded participants with missing major risk factor (hypertension and smoking history) and anthropometric data (n=38). The population for machine learning (ML) analysis included 3,679 participants. For regression analyses, we further excluded participants with missing covariates (n=768). The population for regression analysis included 3,001 participants (Figure 1).

**Figure 1.**
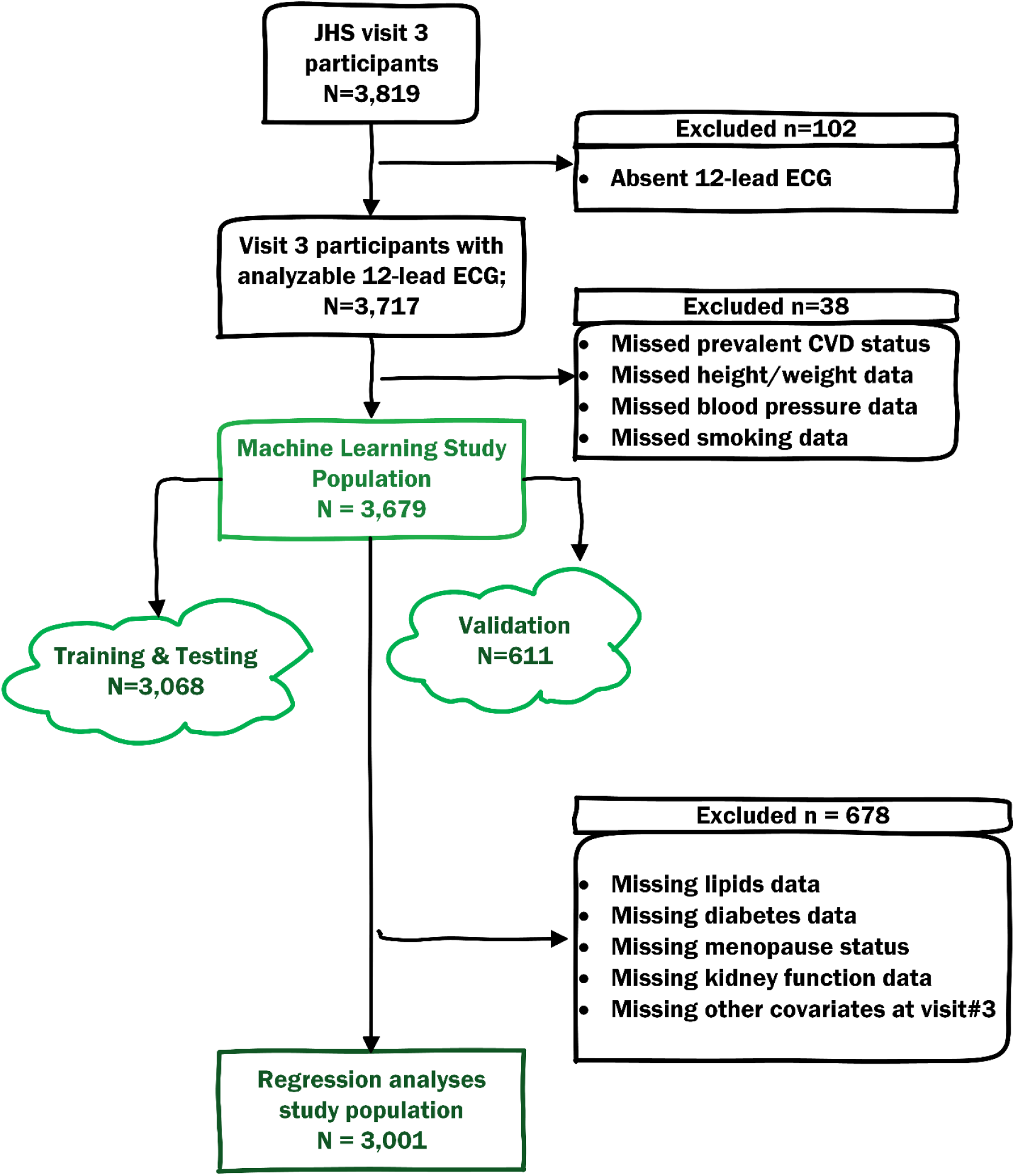
Flowchart of study cohort development.

### ECG and VCG analysis

Raw digital ECG signal was analyzed in the Tereshchenko laboratory at OHSU, as previously described.^14, 15, 23, 24^ Briefly, the analysis includes several steps. First, each cardiac beat was manually labeled by at least two physician investigators (KL, KP, LGT). Then, 12-lead ECG was transformed into XYZ ECG, using Kors transformation.^25^ Using only one (dominant) type of beat, the time-coherent global median beat was constructed, and the origin of the heart vector was identified.^24^ In this study, we included three categories of median beats. Normal (N) category included normal sinus median beat, atrial paced median beat, junctional median beat, and ectopic atrial median beat. The ventricular pacing (VP) category included ventricular paced and both atrial and ventricular paced median beats. The supraventricular (S) category included median beats of atrial fibrillation or atrial flutter with consistently one type of ventricular conduction.

Spatial peak and spatial area QRS, T, and spatial ventricular gradient (SVG) vectors were constructed, and their direction (azimuth and elevation) and magnitudes were measured.^14, 23, 24^ Scalar values of SVG were measured by sum absolute QRST integral (SAI QRST)^26-28^ and by QT integral on vector magnitude (VM) signal.^23^ Both area and peak QRS-T angles were measured.^14, 23, 24^ Quality control of automated ECG analysis was performed by investigators (KTH, NMR) with the aid of visual display. The open-source MATLAB (MathWorks, Natick, MA, USA) code is provided at https://physionet.org/physiotools/geh & https://github.com/Tereshchenkolab/Origin.

Traditional ECG measurements were performed by the 12 SL algorithm as implemented in Magellan ECG Research Workstation V2 (GE Marquette Electronics, Milwaukee, WI) and included median beat measurements (PR, QRS, QT intervals, and frontal P, QRS, and T axes), as well as durations, amplitudes, and areas of all identified by the algorithm waves and segments on all 12 leads. We used the results of automated 12-lead ECG measurements as reported by the 12SL algorithm, without further quality control procedures. QT interval was corrected for heart rate by several approaches: Bazett, Fridericia, Hodge, and Framingham, as provided by the JHS Coordinating Center. Cornell voltage was calculated as the sum of the R_aVL_ and the S_V3_ amplitudes. Frontal QRS-T angle was calculated as previously described.^29^

### Prevalent cardiovascular disease

Prevalent CVD was defined during the 3^rd^ clinical examination if at least one of the following was present: (1) history of coronary heart disease (CHD) defined as either self-reported prior MI (diagnosed by a doctor or health professional, or hospitalization for MI), or ECG diagnosis of MI, (2) history of cardiac procedure defined as either prior coronary revascularization [coronary artery bypass grafting (CABG) or percutaneous coronary intervention (PCI)] or peripheral arterial revascularization, or (3) prior carotid angioplasty or carotid endarterectomy, or (4) self-reported stroke history (diagnosed by a doctor or health professional).

### Covariates: Cardiovascular risk factors measured at the 3_rd_ clinical examination

The 3^rd^ clinical examination included physical examination, anthropometry, a survey of medical history and cardiovascular risk factors, and the collection of blood and urine. Height and weight were measured, and body mass index (BMI) and body surface area (BSA) were calculated. BMI categories included under- or normal weight (<25.0 kg/m^2^), overweight (25.0 to <30.0 kg/m^2^), or obese (≥30.0 kg/m^2^). The dimensionless waist-to-hip ratio (WHR) was calculated as the ratio of the circumference of the waist to that of the hips. Self-reported post-menopausal status for women was defined as no menstrual periods during the past two years.^30^

Smoking status was defined as current, former, and never smoker. The use of alcohol was categorized as yes (in the past 12 months) versus no. Physical activity was characterized according to the American Heart Association (AHA) classification^31^ as ideal (≥75 minutes of vigorous or ≥150 minutes of moderate or combined physical activity per week), intermediate (<75 minutes of vigorous or <150 minutes of moderate or combined physical activity per week), or poor (no vigorous or moderate physical activity).

Hypertension was defined as blood pressure ≥ 140/90 mm Hg or use of antihypertensive therapy. Fasting plasma glucose and glycosylated hemoglobin (HbA1c) levels were measured as previously described.^32^ Diabetes was defined per 2010 American Diabetes Association guidelines as fasting plasma glucose ≥ 126 mg/dl or HbA1c ≥ 6.5% or use of antidiabetic medications within 2 weeks prior to the clinic visit. Fasting total cholesterol, low-density lipoprotein (LDL), high-density lipoprotein (HDL), and triglyceride levels were measured.^33^

The estimated glomerular filtration rate (eGFR_CKD-EPI_) was calculated using the Chronic Kidney Disease Epidemiology Collaboration equation (ml/min/1.73 m^2^). Self-reported history of chronic kidney disease (CKD) and dialysis was recorded. Systemic inflammation was assessed by high sensitivity C-Reactive Protein (hsCRP), which was measured in serum as previously described.^34^

Family income was categorized as at least $75,000 per year versus less than $75,000 per year.

### Families structure

Per the design, the JHS enrolled the secondary family members and comprised a Family Cohort that included nearly 300 pedigrees.^35^ In this study, to assess the effect of unmeasured environmental and genetic factors, we comprised family units of participants with the same 4-symbols code indicating similar family name.

### Statistical analysis

#### Unadjusted comparison

Normally distributed continuous variables were reported as mean ± standard deviation (SD) and compared using the *t*-test. Variables with a skewed distribution were reported as the median and interquartile range (IQR) and compared using the Wilcoxon rank-sum (Mann-Whitney) test. Categorical variables were compared using the χ2 test.

#### Analysis of circular variables

Circular variables (azimuth and elevation angles of QRS, T, and SVG vectors, and QRS-T angles) were presented as mean and 95% confidence interval (CI). Two-sample tests for circular variables included the Watson U-square statistic and the Kuiper statistics.

As distributions of QRS-T angles and SVG elevation angles were normal or nearly normal, and their values were only positive, ranging from 0 to 180 degrees, we included them in all regression analyses without transformation. As SVG azimuth angles were ranging from −180° to +180°, we transformed SVG azimuth by doubling its value and then adding 360°^11, 36^. For interpretation, we then transformed SVG azimuth back.

#### Adjusted Linear Models

To answer whether sex and prevalent CVD are independently associated with GEH (Figure 2A), we constructed linear models with ECG and VCG measurements as outcome variables (one-by-one) and adjusted for known confounders^15, 17, 26, 28, 37, 38^ that were measured during the 3^rd^ clinical examination. All models were adjusted for age, anthropometric characteristics (weight, height, BMI, BSA, waist and hip circumference, WHR), lipid levels (total cholesterol, LDL, HDL, triglycerides), hypertension and levels of systolic and diastolic BP, diabetes and levels of fasting glucose and HbA1c, CKD, history of dialysis, and eGFR_CKD-EPI_, hsCRP, levels of physical activity, smoking, use of alcohol, menopausal state, and socioeconomic factors (income category). To account for unmeasured confounders, we adjusted for the study recruitment type. All models were also adjusted for the type of median beat (N, S, or VP). All models except the model for heart rate, were adjusted for mean RR’ interval. As we previously showed that sex modifies an association of GEH with SCD^12^, we included an interaction term of sex with prevalent CVD status in all models.

**Figure 2.**
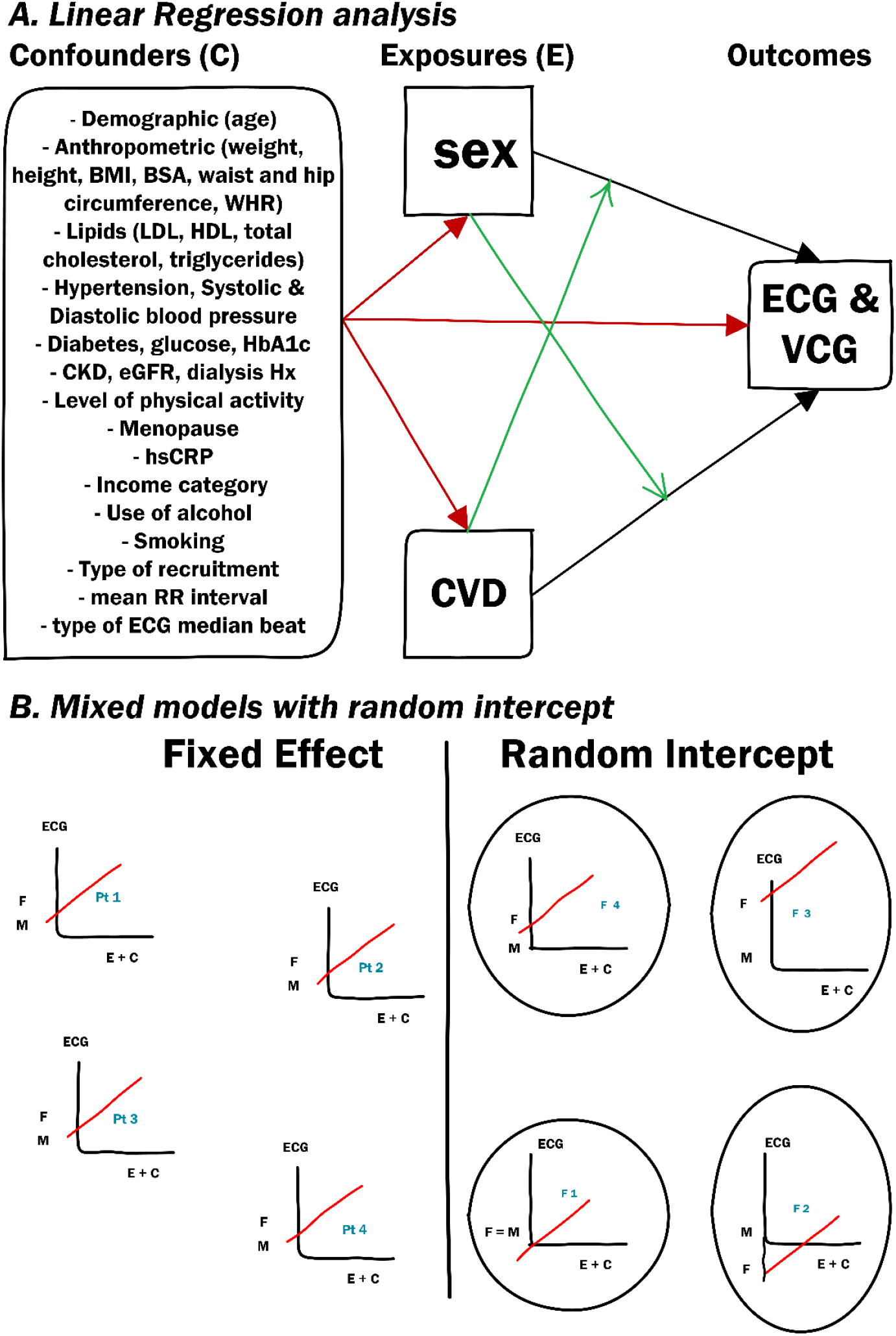
**A**. Directed acyclic graph of the regression analysis. Black arrows indicate the studied associations. Green arrows indicate interaction (effect modification). Red arrows connect confounders with exposures and outcomes. **B**. Schematic illustration of a linear regression model (or fixed “within family” effect) and a mixed model with a random intercept (between families effect).

The linear model assumes that the error terms are independent, which may not be the case in our study, as the JHS enrolled families and the error terms are likely correlated within families. Therefore, we first compared the fit of two models: linear regression and mixed linear model with random intercept (Figure 2B). Measuring a random effect is a way of accounting for unmeasured differences between family units. Intercept is a predicted value of outcome if all predictors in the model are equal to zero (at the reference level). We used the likelihood ratio test to compare the fit of linear regression and mixed linear model with random intercept. If a mixed model with random intercept was a better fit, we further used the generalized least squares (GLS) estimator, which does not require normality of the residuals. GLS is a weighted average of between and within effects. We reported both fixed (within families) and random (between families) effects. We used the Hausman specification test to determine whether we should be allowed to use a GLS estimator, or if we should use the fixed (within) effect model only. The Hausman specification test describes whether there are systematic differences between the GLS and fixed effect estimators due to the correlation of predictor variable with the error term (omitted variable bias, or endogeneity).

#### Machine learning approach for detection of prevalent CVD

We randomly split ML study population into two non-overlapping samples in such a way that each cluster was completely contained within one set: training and testing (80%; 678 families; n=3,068), and validation (20%; 169 families; n=611).

Considering future implementation of our CVD detection tool in AA communities, we included predictor variables that can be easily obtained in barbershops: age, sex, anthropometric characteristics (height, weight, BMI, BMI categories, BSA), history of hypertension, systolic and diastolic BP, smoking history, and automatically measured ECG and VCG metrics (43 variables, listed in Supplemental Table 3).

We fitted 8 different models (random forests^39^, convolutional neural network^40^, lasso, adaptive lasso, plug-in-based lasso, elastic net, ridge with penalized and post-selection coefficients, and logistic regression).

To train the random forest algorithm, we arranged the data in a randomly sorted order, and tuned the number of subtrees and number of variables to randomly investigate at each split. We used both out-of-bag error (tested against training data subsets that are not included in subtree construction) and a validation error (tested against the validation data) to find the model with the highest testing accuracy.

We trained the convolutional neural network with 20 hidden layers, using 500 iterations with a training factor 2, and 4 normalization parameters. For the model with VCG input (43 variables), the network was comprised of 3 layers, 64 neurons per layer, and 901 synapse weights. For the model with ECG input (153 variables), the network was comprised of 3 layers, 174 neurones per layer, and 3,101 synapse weights.

The least absolute shrinkage and selection operator (lasso) family of models utilized ten-fold cross-validation in the training sample. In lasso model, cross-validation selected the tuning parameter λ that minimized the out-of-sample deviance. The adaptive lasso performed a multistep cross-validation, performing second cross-validation step among the covariates selected in the first cross-validation step. The plug-in-based lasso used partialing-out estimators to determine which covariates belong in the model, achieving an optimal bound on the number of covariates it included.^41^ The elastic net is an extension of the lasso that permits retention of correlated covariates.^42^ In the ridge model, the penalty parameter used squared terms and kept all predictors in the model.

We compared the predictive accuracy of the models by comparing the area under the receiver operator curve (ROC AUC). As the goal of screening is to identify all individuals with prevalent CVD, we strived to maximize the sensitivity of the test, and we selected a 100% sensitivity threshold. We validated the CVD detection tool in the validation sample by measuring ROC AUC and assessing the sensitivity and specificity of the selected at the previous step threshold.

To assess calibration, we evaluated the goodness of fit in validation sample, using several approaches. We compared the observed and predicted proportions within the groups formed by the Hosmer-Lemeshow test.^43^ We also used the calibration belt^44^ to examine the relationship between estimated probabilities and observed CVD rates. For the lasso family of models, we also calculated out-of-sample deviance and deviance ratio.

#### Comparison of machine learning models using the input of 12SL algorithm ECG features

To compare the composition and predictive accuracy of ML models using ECG features measured on 12-lead ECG by the 12SL algorithm (GE Marquette Electronics, Milwaukee, WI), we repeated the described above ML steps with the input of additional 652 variables, which included frontal QRS-T angle and fine 12-lead ECG features (amplitudes, durations, and areas of all ECG waves). We compared random forests, convolutional neural networks, lasso, adaptive lasso, plug-in-based lasso, and elastic net with penalized and post-selection coefficients. Due to a large number of input variables (n=695), we did not test the performance of logistic regression and ridge models as nearly all the predictors were kept in the model. For an adequate comparison of convolutional neural networks with VCG and ECG input, a model with ECG input did not include VCG variables, and included only measurements of main ECG waves, without “prime” ECG waveforms (153 variables).

Statistical analysis was performed using STATA MP 16.1 (StataCorp LP, College Station, TX). *P*-value < 0.05 was considered statistically significant. STATA code is provided at https://github.com/Tereshchenkolab/statistics.

## Results

### Study population

On average, study participants were 62 years of age; more than half were female (Table 1) and were obese. Nearly three-quarters of participants had hypertension, and one-third of the participants were current or former smokers. Prevalent CVD was diagnosed in 411 out of 3,679 participants (11.2%).

**Table 1.** Comparison of clinical characteristics in men and women

| Characteristic | All (n=3,001) | Men (n=1,151) | Women (n=1,850) | P-value |
| --- | --- | --- | --- | --- |
| Age(SD), y | 62.4(11.5) | 60.7(11.8) | 63.5(11.1) | <0.0001 |
| BMI(SD), kg/m<sup>2</sup> | 32.0(6.3) | 30.4(6.3) | 32.9(7.3) | <0.0001 |
| Obese BMI group, n(%) | 1,677(55.9) | 524(45.5) | 1,153(62.3) | <0.0001 |
| Waist-hip ratio(SD) | 0.91(0.08) | 0.96(0.06) | 0.89(0.07) | <0.0001 |
| BSA(SD), m<sup>2</sup> | 2.00(0.24) | 2.12(0.23) | 1.93(0.21) | <0.0001 |
| Ever tobacco smoker, n(%) | 906(30.2) | 470(40.8) | 436(23.6) | <0.0001 |
| Alcohol intake past 12mo, n(%) | 1,356(45.2) | 660(57.3) | 696(37.6) | <0.0001 |
| Hypertension, n(%) | 2,225(74.1) | 790(68.6) | 1,435(77.6) | <0.0001 |
| Systolic blood pressure(SD), mmHg | 127.9(18.5) | 128.5(16.9) | 127.6(19.4) | 0.153 |
| Diastolic blood pressure (SD), mmHg | 75.2(10.7) | 76.6(10.8) | 74.3(10.5) | <0.0001 |
| HbA1c(SD), % | 6.13(1.10) | 6.09(1.14) | 6.16(1.07) | 0.130 |
| Plasma glucose(SD), mg/dL | 105.5(32.3) | 107.6(34.3) | 104.3(30.9) | 0.008 |
| Diabetes status, n(%) | 881(29.4) | 315(27.4) | 566(30.6) | 0.059 |
| LDL cholesterol(SD), mg/dL | 120.3(36.2) | 119.7(36.4) | 120.7(36.1) | 0.488 |
| HDL cholesterol(SD), mg/dL | 57.9(16.0) | 52.5(14.7) | 61.3(15.9) | <0.0001 |
| Triglyceride(SD), mg/dL | 97.4(48.7) | 102.5(54.2) | 94.2(44.7) | <0.0001 |
| Total cholesterol(SD), mg/dL | 197.7(40.1) | 192.7(40.1) | 200.8(39.7) | <0.0001 |
| Stroke history, n(%) | 155(5.2) | 63(5.5) | 92(5.0) | 0.547 |
| CHD history, n(%) | 104(3.5) | 48(4.2) | 56(3.0) | 0.096 |
| Cardiac procedures history, n(%) | 98(3.3) | 37(3.2) | 61(3.3) | 0.905 |
| CVD history, n(%) | 311(10.4) | 132(11.5) | 179(9.7) | 0.117 |
| Menopause, n(%) | - | - | 1,688(91.2) | - |
| High sensitivity CRP median(IQR), mg/dL | 0.29(0.12-0.64) | 0.20(0.09-0.43) | 0.36(0.15-0.78) | <0.0001 |
| eGFR CKD-EPI | 85.7(22.2) | 85.1(21.7) | 86.1(22.5) | 0.215 |
| Dialysis history, n(%) | 22(0.7) | 12(1.0) | 10(0.5) | 0.117 |
| CKD History, n(%) | 91(3.0) | 30(2.6) | 61(3.3) | 0.283 |
| Ideal Physical activity, n(%) | 764(25.5) | 361(31.4) | 403(21.8) | <0.0001 |
| Income ≥75K/year, n(%) | 623(20.8) | 364(31.6) | 259(14.0) | <0.0001 |
| Heart rate(SD), bpm | 63.6(10.5) | 62.8(10.6) | 64.1(10.4) | 0.002 |
| QRS duration(SD), ms | 89.2(15.7) | 93.6(16.3) | 86.4(14.6) | <0.0001 |
| Bazett corrected QT (SD), ms | 426.5(25.5) | 418.3(26.5) | 431.5(23.5) | <0.0001 |
| Framingham corrected QT (SD),ms | 422.2(22.6) | 414.8(23.3) | 426.9(20.8) | <0.0001 |
| Hodge corrected QT (SD), ms | 423.6(22.9) | 416.9(23.6) | 427.8(21.4) | <0.0001 |
| Fridericia corrected QT (SD), ms | 423.1(22.6) | 415.8(23.3) | 427.6(20.9) | <0.0001 |
| Cornell voltage(SD), μV | 1534(592) | 1706(650) | 1426(525) | <0.0001 |
| Median beat: normal sinus, n(%) | 2,957(98.5) | 1,133(98.4) | 1,824(98.6) | 0.093 |
| Median beat: atrial fibrillation, n(%) | 32(1.1) | 10(0.9) | 22(1.2) |  |
| Median beat: ventricular pacing, n(%) | 12(0.4) | 8(0.7) | 4(0.2) |  |
| QRS area(SD), mV*ms | 38.5(18.3) | 39.5(20.0) | 37.9(17.2) | 0.025 |
| Peak QRS magnitude(SD), mV | 1.59(0.44) | 1.60(0.46) | 1.59(0.42) | 0.540 |
| Area QRS azimuth(95%CI),° | 20.7(19.9-21.5) | 21.4(19.8-23.0) | 20.3(19.3-21.2) | <0.001 |
| Peak QRS azimuth(95%CI),° | 9.3(8.5-10.1) | 8.3(6.7-9.9) | 9.9(9.0-10.8) | <0.001 |
| Area QRS elevation(95%CI),° | 73.8(73.2-74.4) | 75.7(74.6-76.8) | 72.6(71.8-73.3) | <0.001 |
| Peak QRS elevation(95%CI),° | 72.7(72.2-73.2) | 74.7(73.8-75.5) | 71.5(70.9-72.1) | <0.001 |
| T area(SD), mV*ms | 48.7(23.3) | 56.5(24.6) | 43.8(21.2) | <0.0001 |
| Peak T magnitude(SD), mV | 0.36(0.16) | 0.40(0.17) | 0.33(0.15) | <0.0001 |
| Area T azimuth(95%CI),° | -45.2(-46.2 to-44.1) | -52.3(-53.9 to-50.7) | -40.6(-41.9 to-39.2) | <0.001 |
| Peak T azimuth(95%CI),° | -36.3(-37.4 to-35.1) | -46.1(-47.8 to-44.4) | -30.0(-31.4 to-28.6) | <0.001 |
| Area T elevation(95%CI),° | 75.8(75.3-76.3) | 77.7(76.9-78.4) | 74.6(73.9-75.3) | <0.001 |
| Peak T elevation(95%CI),° | 70.1(69.5-70.6) | 73.3(72.5-74.1) | 68.0(67.3-68.7) | <0.001 |
| Wilson's (area) SVG (SD), mV*ms | 69.6(28.5) | 73.7(31.1) | 67.0(26.3) | <0.0001 |
| Peak SVG magnitude (SD), mV | 1.81(0.50) | 1.81(0.54) | 1.81(0.47) | 0.860 |
| Area SVG azimuth (95%CI),° | -14.3(-15.1 to-13.4) | -22.2(-23.6 to-20.9) | -9.4(-10.4 to-8.5) | <0.001 |
| Peak SVG azimuth (95%CI),° | 3.4(2.6-4.1) | 0.6(-0.9 to 2.1) | 5.0(4.2-5.8) | <0.001 |
| Area SVG elevation (95%CI),° | 71.8(71.3-72.3) | 74.0(73.1-74.9) | 70.4(69.8-71.1) | <0.001 |
| Peak SVG elevation (95%CI),° | 71.2(70.7-71.6) | 73.1(72.3-73.9) | 69.9(69.4-70.5) | <0.001 |
| SAI QRST(SD), mV*ms | 154.5(51.7) | 169.4(55.4) | 109.1(46.9) | <0.0001 |
| VM QT integral (SD), mV*ms | 103.4(34.5) | 113.8(36.9) | 97.0(31.3) | <0.0001 |
| Area QRS-T angle(95%CI),° | 67.3(66.1-68.6) | 75.0(72.9-77.1) | 62.7(61.2-64.2) | <0.001 |
| Peak QRS-T angle(95%CI),° | 48.3(47.1-49.6) | 55.7(53.4-57.9) | 44.1(42.7-45.5) | <0.001 |

VM=vector magnitude

There were few differences in the characteristics of participants with missing covariates who were excluded from the regression analyses. Excluded participants were more likely to be younger females, with smaller height, lower systolic blood pressure, higher BMI, and faster heart rate (Supplemental Table 1). Nevertheless, VCG characteristics of included and excluded participants were broadly similar.

### Family units structure

There were 863 family units in our study. Nearly half of them consisted of a single person (343 units; 40%), and 17% (149 units) consisted of two participants. There were 16 large family units (2%) with 20-79 family members per unit, accounting for 24% of the study population (n=713).

For the vast majority of linear models, the linear regression model provided a better fit than the mixed model. Only four mixed models with random intercept demonstrated better fit than linear regression: models for QRS duration, area and peak T azimuth, and area SVG azimuth. Hausman specification test supported the use of the GLS estimator in these four models.

### Comparison of men and women

Female study participants were older, less physically active, with a higher prevalence of obesity and hypertension, higher levels of hsCRP, and a lower income than male participants (Table 1). On the other hand, women were less likely to smoke and consume alcohol and had a more favorable lipid profile than men. There were no statistically significant differences in the CVD prevalence between men and women (Table 1).

In unadjusted comparison (Table 1), women had a faster heart rate, more narrow QRS, and longer QTc than men. There were no differences in peak SVG and peak QRS magnitudes between men and women; however, SAI QRST and Wilson’s (area) SVG, as well as QRS-T angles were smaller in women than in men. There were significant differences in SVG direction: SVG pointed higher up and further anteriorly in men than in women. There were no differences in the type of median beat between men and women; only approximately 1% of participants had S and VP types of the median beat (Table 1).

After adjustment for confounders (Table 2), the QRS-T angle remained larger in men than in women (Figure 3). The SVG vector pointed farther upward and anteriorly in men than in women (Figure 4). A significant random effect for area SVG azimuth (Figure 5) indicated a range of meaningful differences (up to 40 degrees) in SVG azimuth for different families. Differences between families in SVG azimuth were mostly due to differences in T azimuth. Both area and peak T vectors pointed more posteriorly in women as compared to men (Figure 6), and there was a significant random effect (Table 2 and Figure 6), resulting in larger sex differences in T azimuth in some families. Wilson’s SVG, SAI QRST, and T area with T vector magnitudes (Figure 7) and elevation of all vectors (Figure 8) were smaller in women than in men. However, there were no sex differences in peak SVG magnitude, and QRS vector azimuth and magnitude (Figures 9-10).

**Figure 3.**
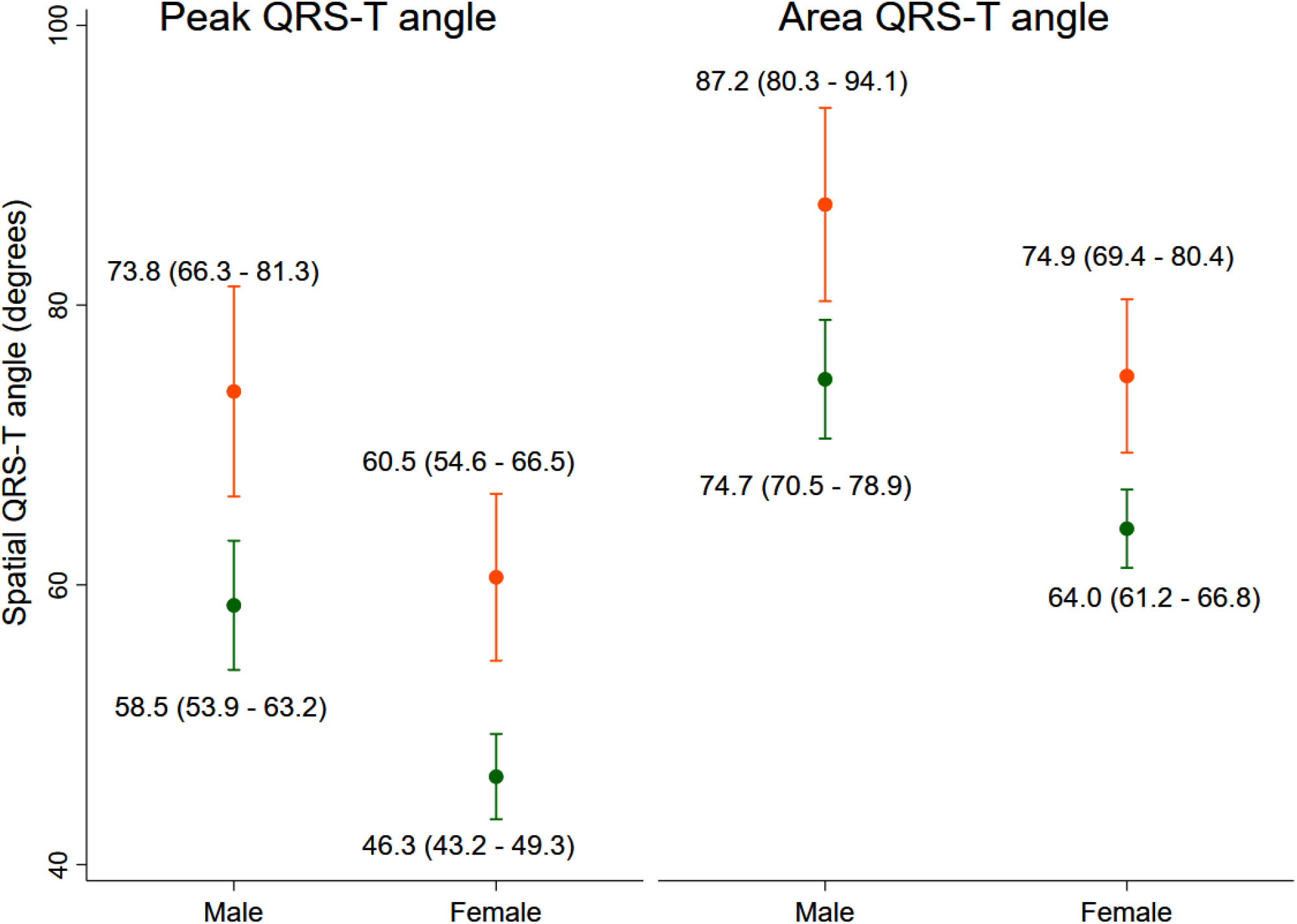
Estimated adjusted marginal (least-squares) means and 95% Confidence Intervals (CI) of peak and area QRS-T angle in male and female participants with (orange) and without (green) prevalent CVD. All models were adjusted for age, weight, height, BMI, BSA, waist and hip circumference, WHR, total cholesterol, LDL, HDL, triglycerides, hypertension, levels of systolic and diastolic BP, diabetes, levels of fasting glucose and HbA1c, CKD, history of dialysis, eGFR_CKD-EPI_, levels of physical activity, smoking, use of alcohol, menopausal state, income, study recruitment, type of median beat, and mean RR’ interval.

**Figure 4.**
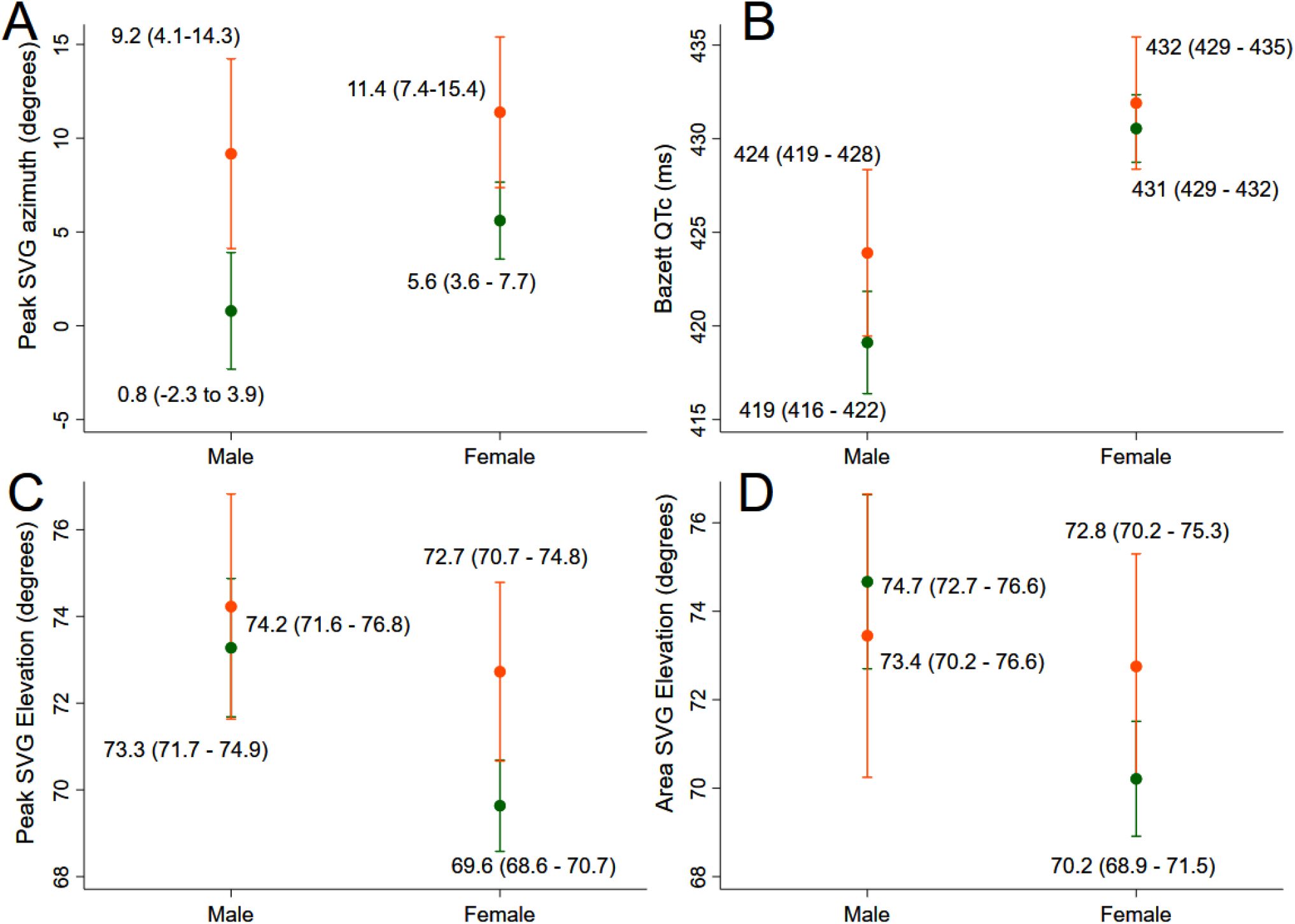
Estimated adjusted (model as described in Figure 3 legend) marginal (least-squares) means and 95% CI of (**A**) peak SVG azimuth, **(B**) Bazett-corrected QT interval, (**C**) peak SVG elevation, (**D**) area SVG elevation in male and female participants with (orange) and without (green) prevalent CVD.

**Figure 5.**
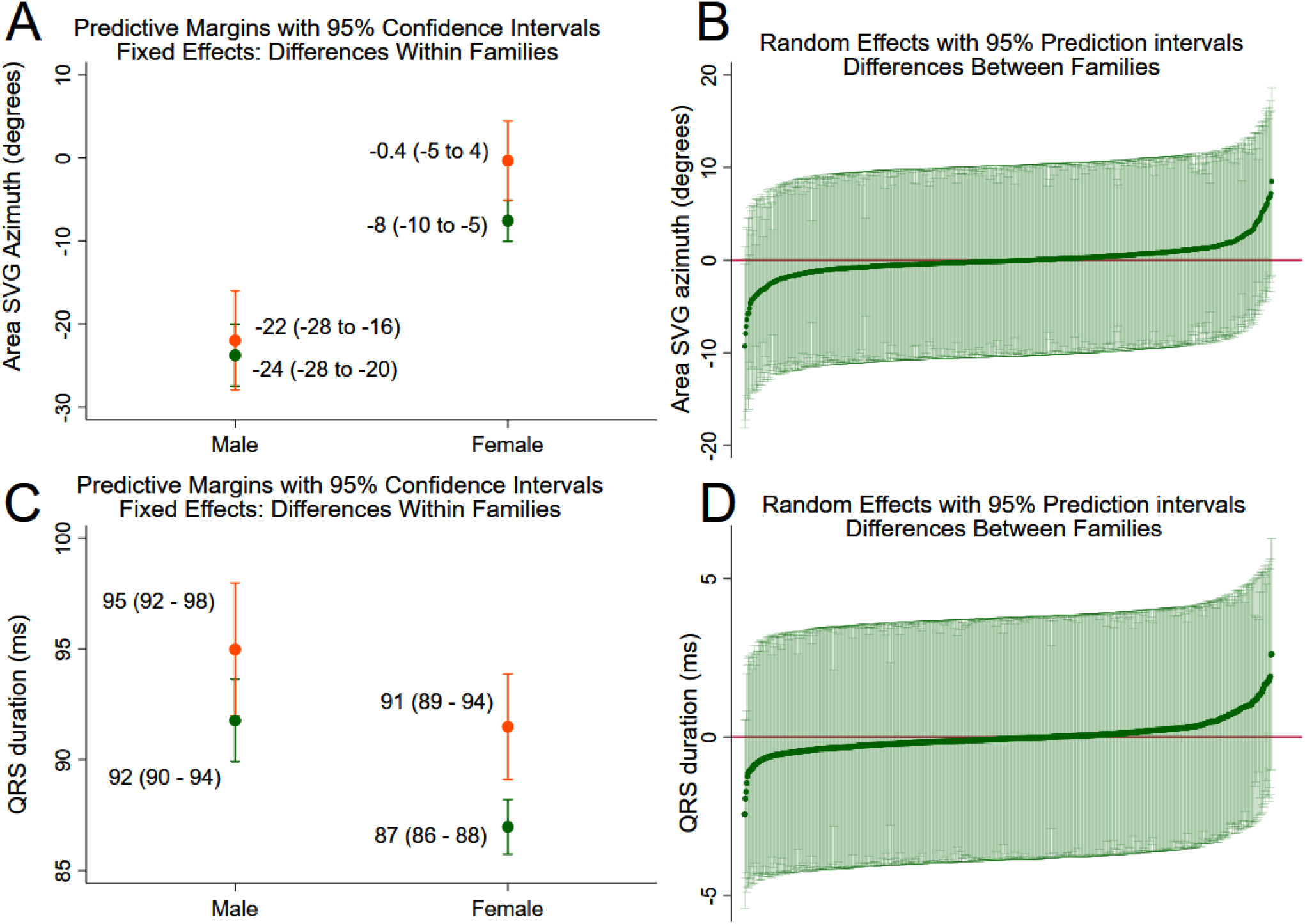
Estimated adjusted (model as described in Figure 3 legend) marginal means and 95% prediction intervals of (**A, B**) area SVG azimuth and (**C, D**) QRS duration. (**A, C**) A fixed portion of a linear prediction (within families effect) in male and female participants with (orange) and without (green) prevalent CVD. (**B, D**) Random intercepts by family (between families effect).

**Figure 6.**
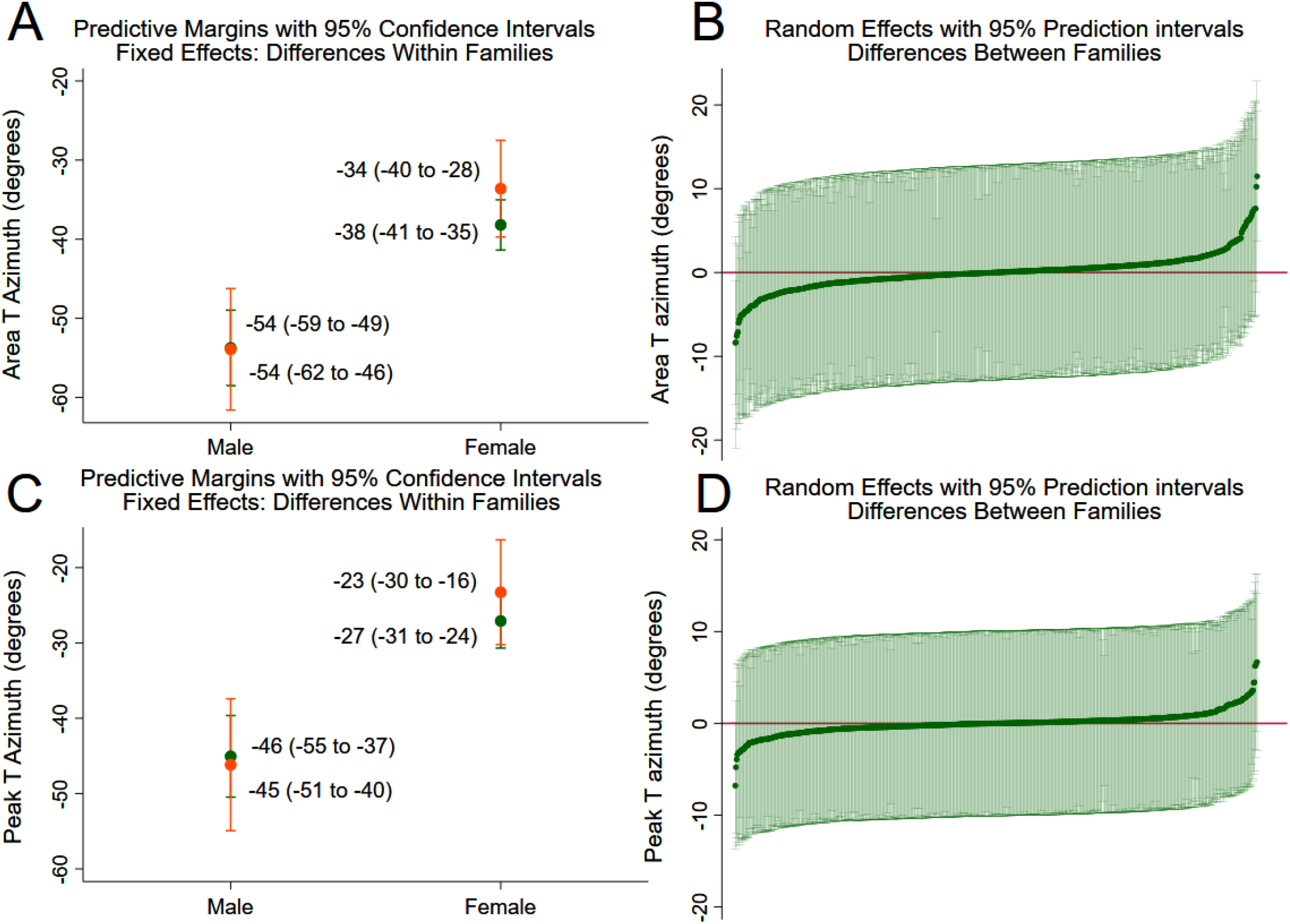
Estimated adjusted (model as described in Figure 3 legend) marginal means and 95% prediction intervals of (**A, B**) area T azimuth and (**C, D**) peak T azimuth. (**A, C**) A fixed portion of a linear prediction (within families effect) in male and female participants with (orange) and without (green) prevalent CVD. (**B, D**) Random intercepts by family (between families effect).

**Figure 7.**
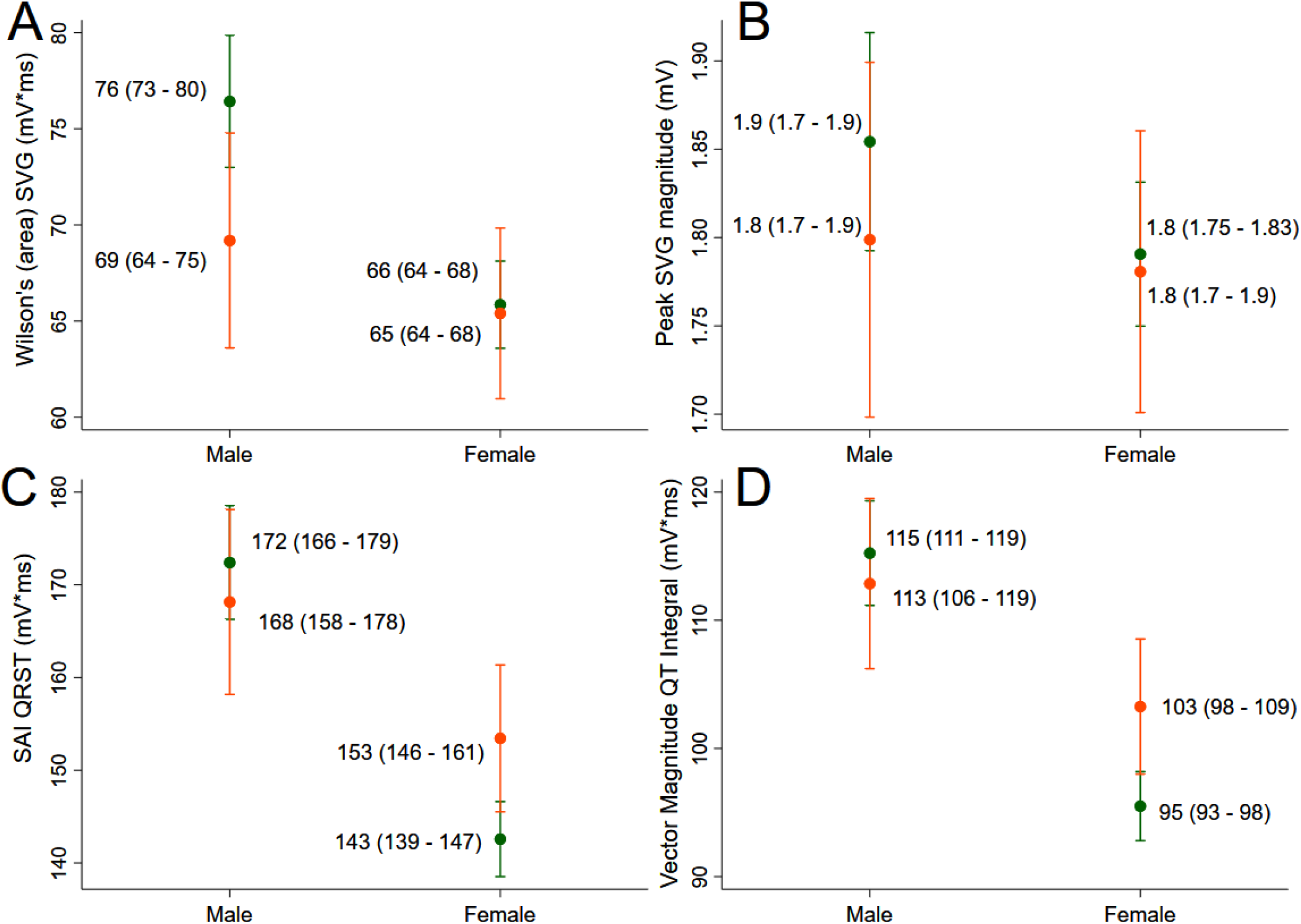
Estimated adjusted (model as described in Figure 3 legend) marginal (least-squares) means and 95% CI of (**A**) Wilson’s (area) SVG, (**B**) peak SVG magnitude, (**C**) SAI QRST, (**D**) vector magnitude QT integral in male and female participants with (orange) and without (green) prevalent CVD.

**Figure 8.**
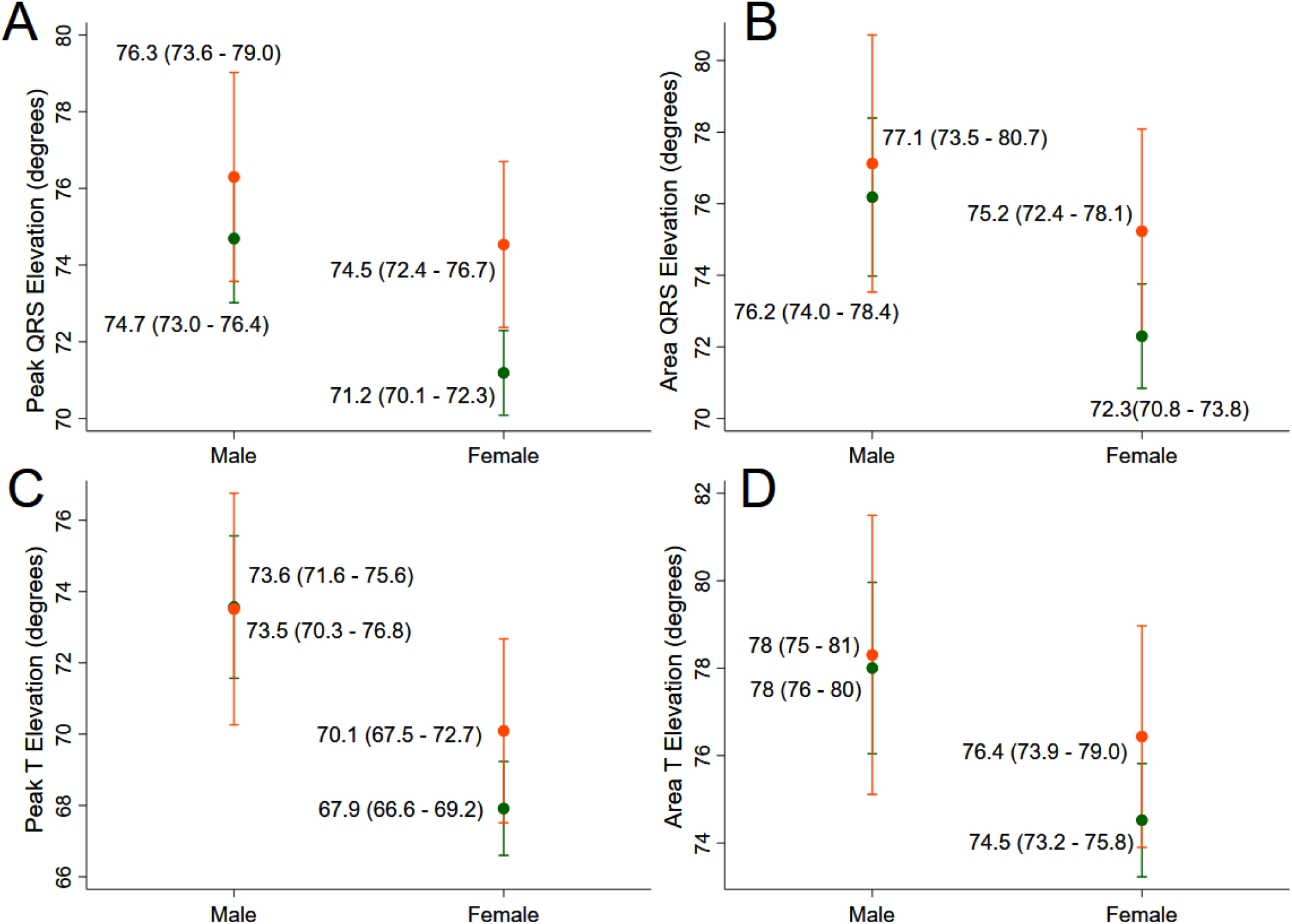
Estimated adjusted (model as described in Figure 3 legend) marginal (least-squares) means and 95% CI of (**A**) peak QRS elevation, (**B**) area QRS elevation, (**C**) peak T elevation, (**D**) area T elevation in male and female participants with (orange) and without (green) prevalent CVD.

**Figure 9.**
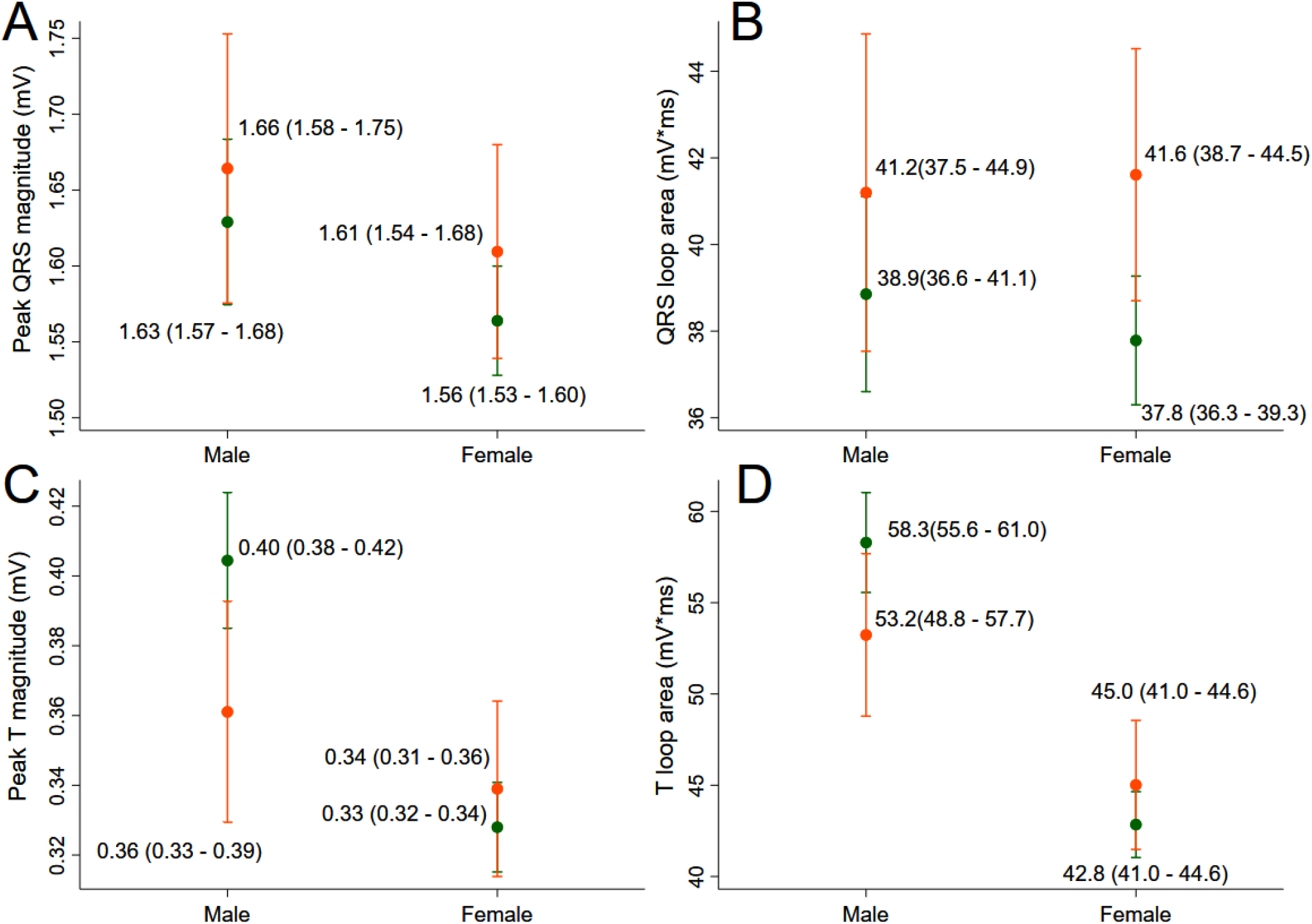
Estimated adjusted (model as described in Figure 3 legend) marginal (least-squares) means and 95% CI of (**A**) peak QRS magnitude, (**B**) QRS area, (**C**) peak T magnitude, (**D**) T area in male and female participants with (orange) and without (green) prevalent CVD.

**Figure 10.**
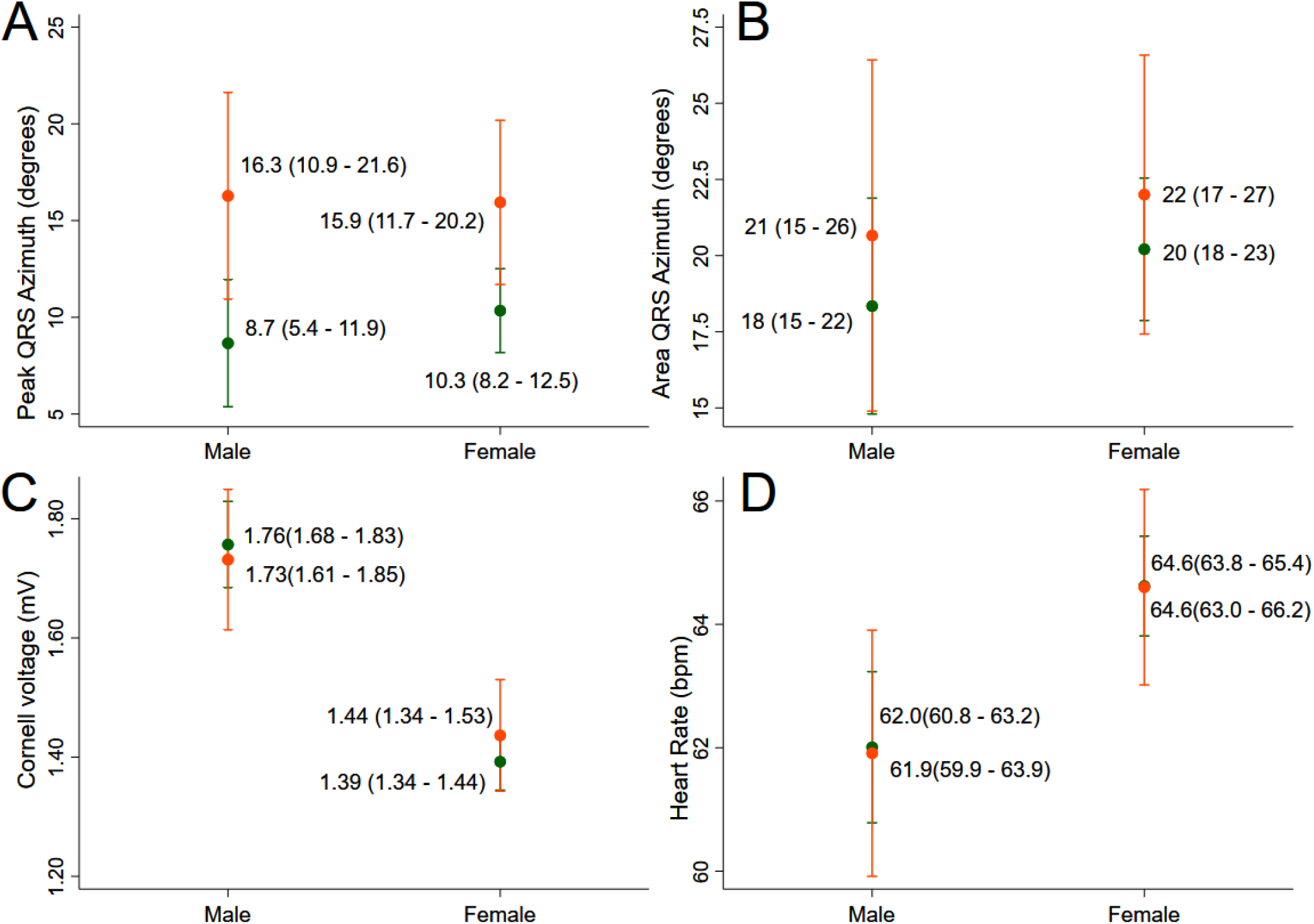
Estimated adjusted (model as described in Figure 3 legend, except model for heart rate, which was not adjusted for RR’ interval) marginal (least-squares) means and 95% CI of (**A**) peak QRS azimuth, (**B**) area QRS azimuth, (**C**) Cornell voltage, (**D**) heart rate in male and female participants with (orange) and without (green) prevalent CVD.

**Table 2.**
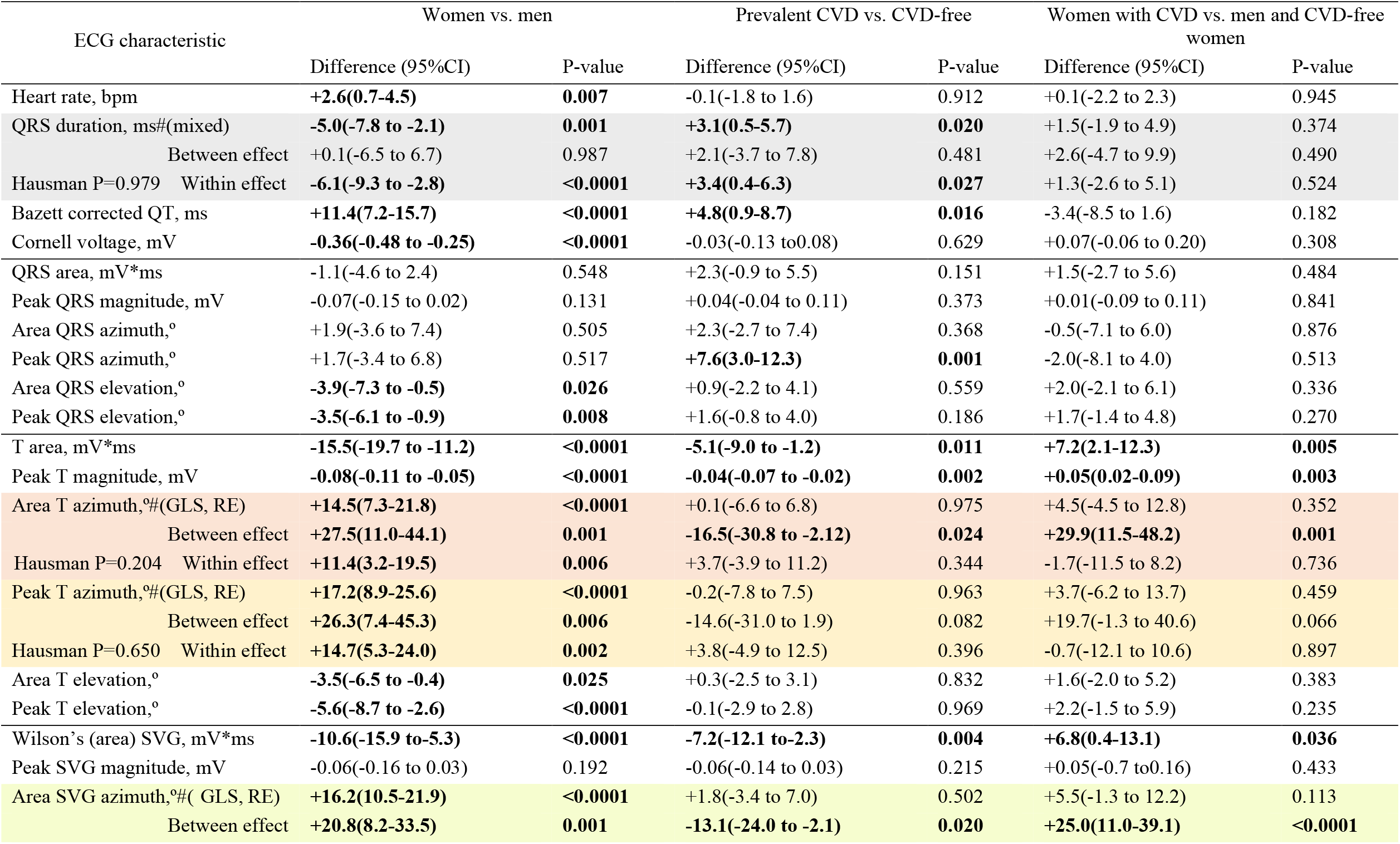
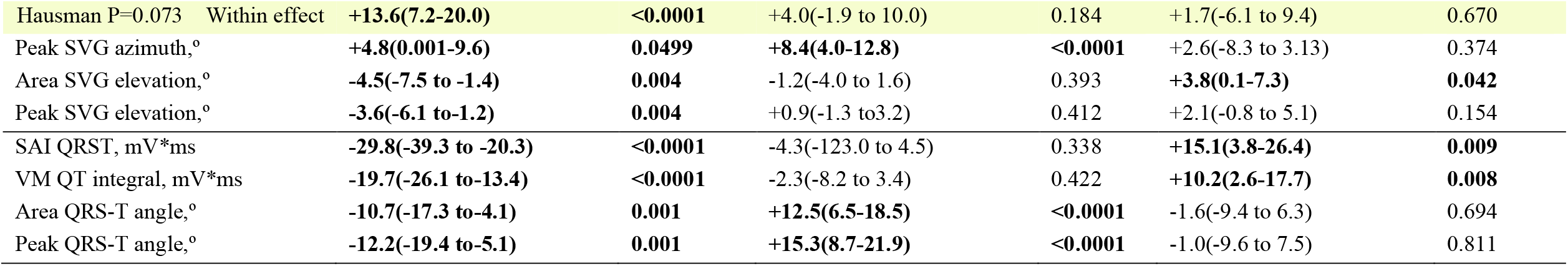
Difference in GEH in women (as compared to men) and participants with prevalent CVD (as compared to CVD-free)

### GEH in participants with and without prevalent CVD

After full adjustment, the QRS-T angle was significantly wider in participants with CVD in both men and women (Figure 3). We observed significant effect modification by sex for several GEH characteristics (Table 2). Women with CVD had larger SAI QRST [by 10.9 (95%CI 3.4-18.3) mV*ms] and VM QT integral [by 7.8 (95%CI 2.8-12.7) mV*ms] than CVD-free women, but there were no differences in men (Figure 7). Men with CVD had smaller Wilson’s SVG [by 7.2 (95%CI 2.3-12.1) mV*ms], T area [by 5.1 (95%CI 1.2-9.0) mV*ms], and T peak magnitude [by 44 (95%CI 16-71) µV] than CVD-free men, whereas no differences by CVD status were observed in women (Figures 7 and 9). In women with CVD, the SVG vector pointed more superiorly [area SVG elevation larger by 2.5 (95%CI 0.2-4.9)°], as compared to CVD-free women (Figure 4). However, there was no difference in SVG elevation in men with and without CVD.

### Development and validation of prevalent CVD detection tool

Training and testing, and validation subsamples were balanced, without major differences in clinical and ECG characteristics between subsamples (Supplemental Table 2).

In tuning the random forest algorithm, we observed that both out-of-bag error and validation error stabilized after 200 iterations at 11-12% (Figure 11), and we conservatively chose 500 subtrees. The minimum validation error (12%) was observed for 23 variables (Figure 23). Thus, we chose 23 variables to randomly investigate at each split. The final random forest model reported small error in validation sample (12.2% or 75 out of 611 individuals), indicating good prediction. However, while the random forest model accurately predicted freedom from CVD in 534 out of 536 participants (specificity 99.6%), it correctly predicted CVD in only 2 out of 75 individuals (sensitivity 2.7%), indicating no clinical usefulness (if used alone). Validation ROC AUC was non-significant (0.512; 95%CI 0.493-0.530). The single most important predictor was sex (Figure 12), which, together with well-known clinical CVD risk factors (age, weight, height, BMI category) comprised the five most important predictors. ECG characteristics had very little impact in the random forest decision tree.

**Figure 11.**
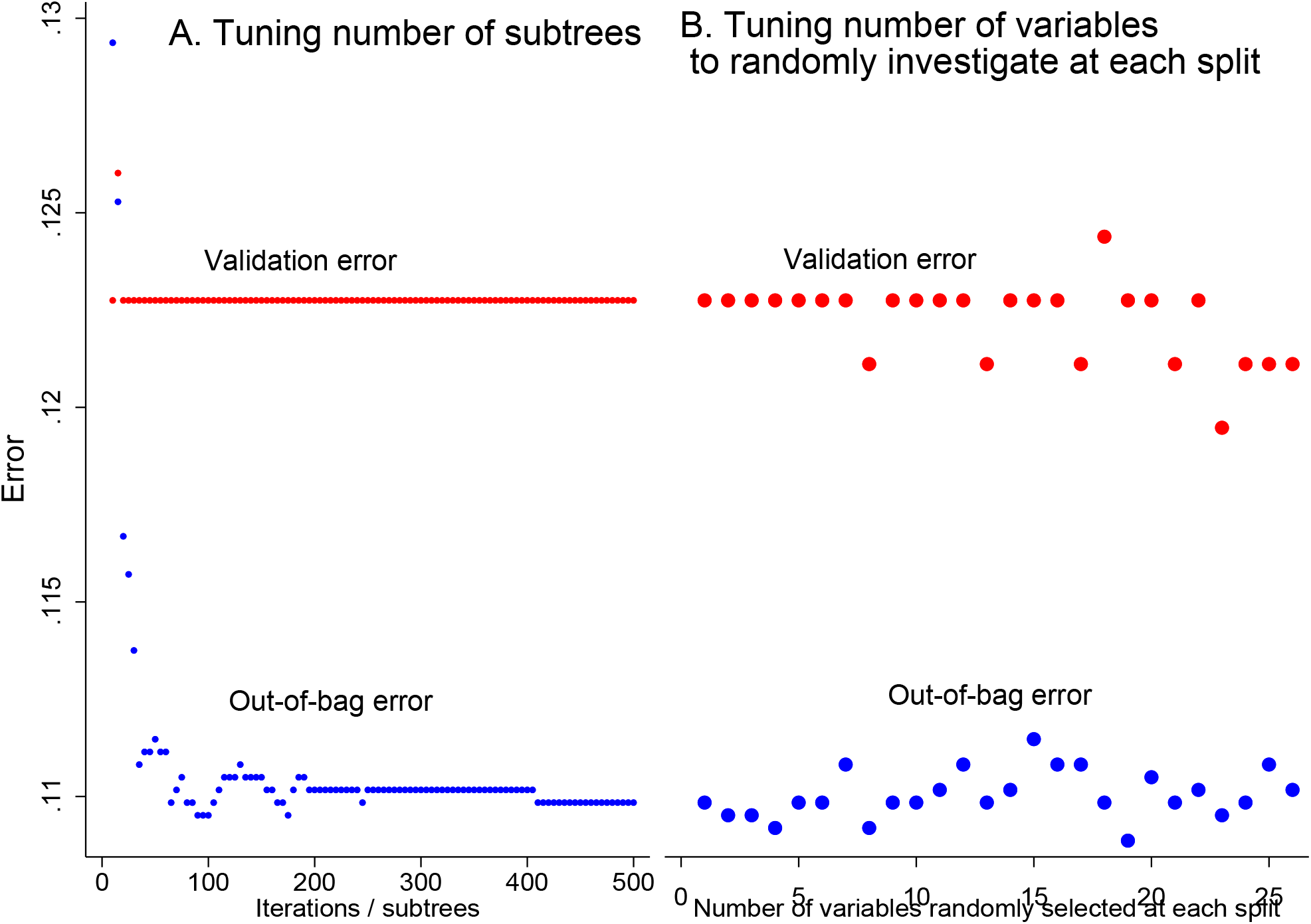
Out-of-bag error and validation error plotted versus (**A**) number of iterations or subtrees, and (**B**) number of variables randomly investigated at each split in a random forest model.

**Figure 12.**
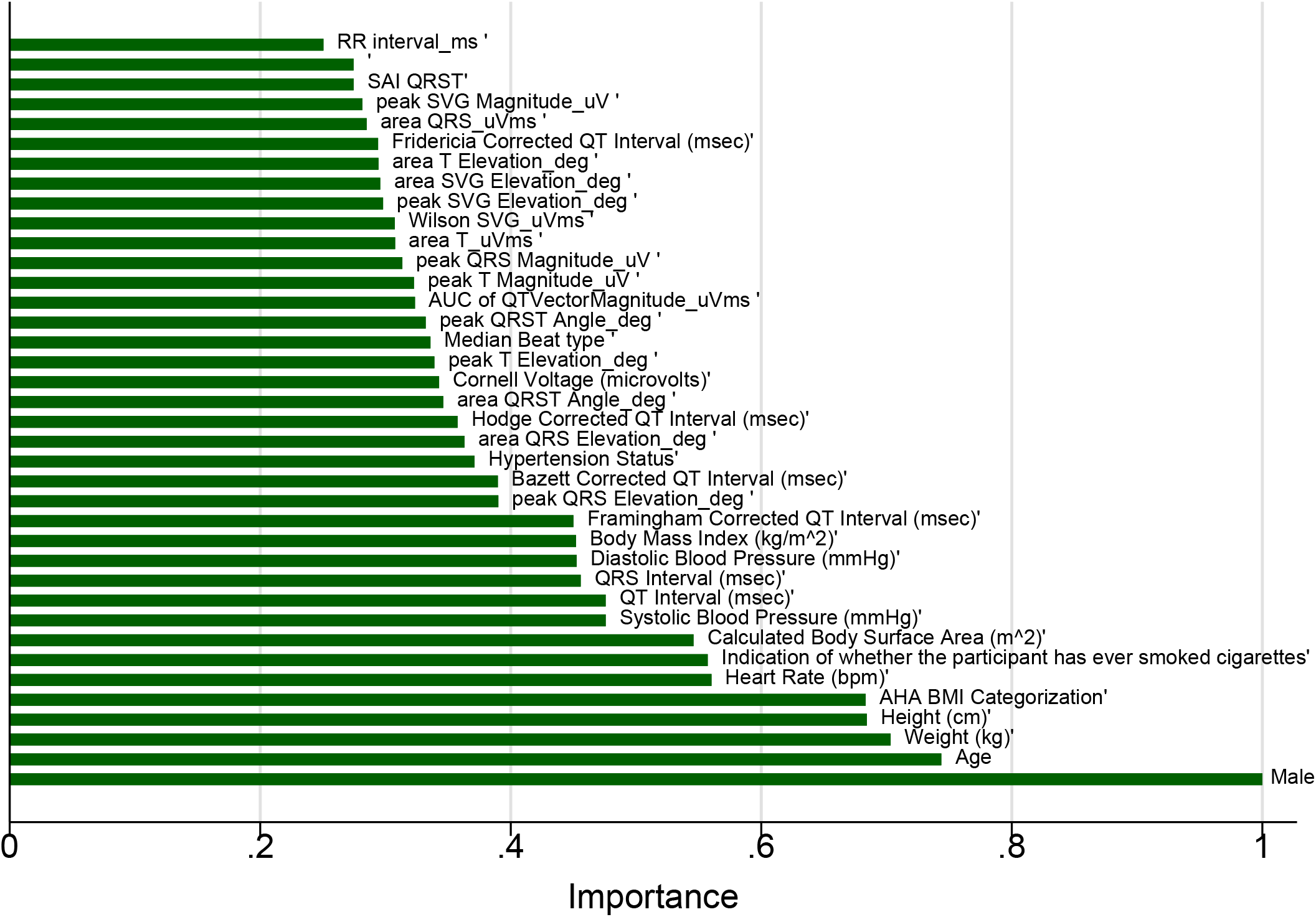
Importance scores of predictor variables in a random forest model with VCG input.

A comparison of the prediction models’ performance is shown in Table 3. Across all models, the convolutional neural network demonstrated the highest predictive accuracy in the training and testing sample, with final error of only 8%. However, the calibration of convolutional neural network model was unsatisfactory (Hosmer-Lemeshow test P<0.0001; Figure 13A and Table 4). Peak QRS-T angle and age demonstrated the largest marginal effect in the convolutional neural network with VCG input (Figure 14).

**Table 3.**
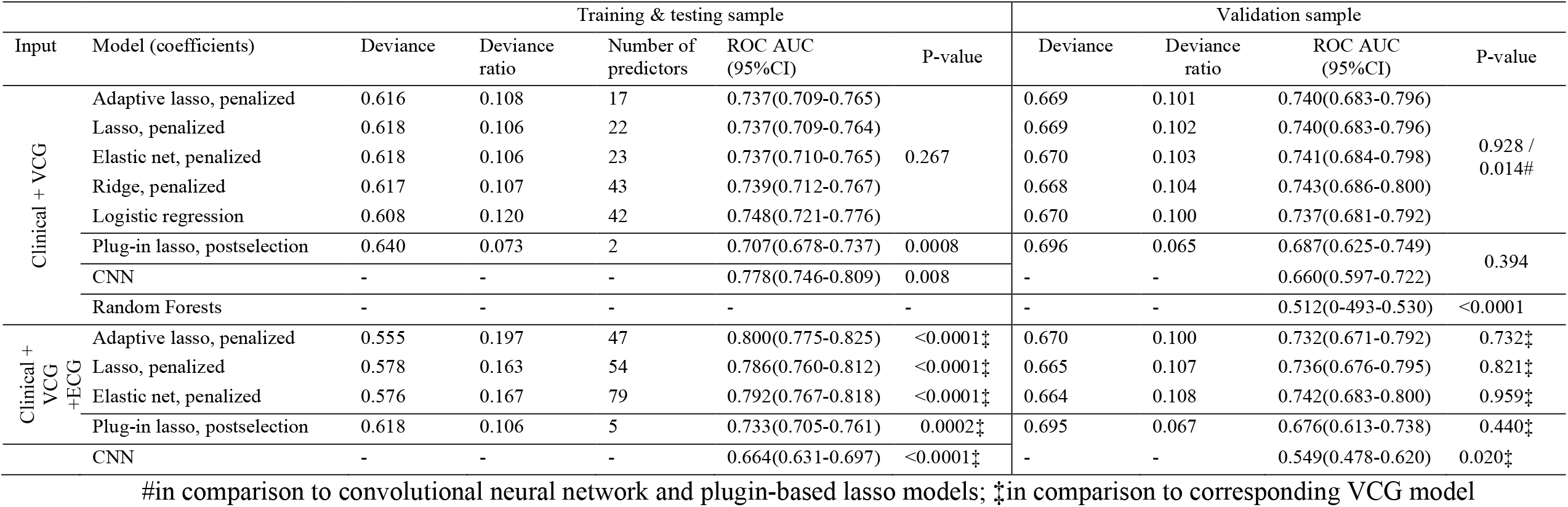
Comparison of models for prevalent CVD detection

**Table 4.** CNN - predicted and observed CVD in deciles of predicted CVD risk

| CVD risk group | N | Observed(%) | Predicted(%) | Min% | Max% | HL $\chi^2$ |
| --- | --- | --- | --- | --- | --- | --- |
| 1 | 3456 | 241(7.0) | 28.1(0.8) | 0 | 9.9 | 1616.05 |
| 2 | 32 | 17(53.1) | 4.4(13.8) | 10.2 | 19.2 | 3.25 |
| 3 | 22 | 9(40.9) | 5.4(24.4) | 20.5 | 28.8 | 4.03 |
| 4 | 17 | 10(58.8) | 6.0(35.5) | 30.9 | 39.8 | 2.51 |
| 5 | 10 | 7(70.0) | 4.5(45.1) | 41.2 | 47.3 | 4.72 |
| 6 | 14 | 12(85.7) | 8.0(57.0) | 50.2 | 60.0 | 1.40 |
| 7 | 26 | 20(76.9) | 17.1(65.9) | 60.9 | 70.0 | 0.38 |
| 8 | 27 | 21(77.8) | 19.6(72.5) | 70.6 | 79.9 | 0.87 |
| 9 | 5 | 5(100) | 4.3(85.2) | 82.7 | 89.5 | 0 |
| 10 | 69 | 68(98.6) | 68.0(98.5) | 91.2 | 100.0 | 1674.92 |
| Total | 3679 | 411(11.2) | 166.5(4.5) | 0 | 100 | 1674.9 |

CNN=convolutional neural network; HL= Hosmer-Lemeshow $\chi^2$

**Figure 13.**
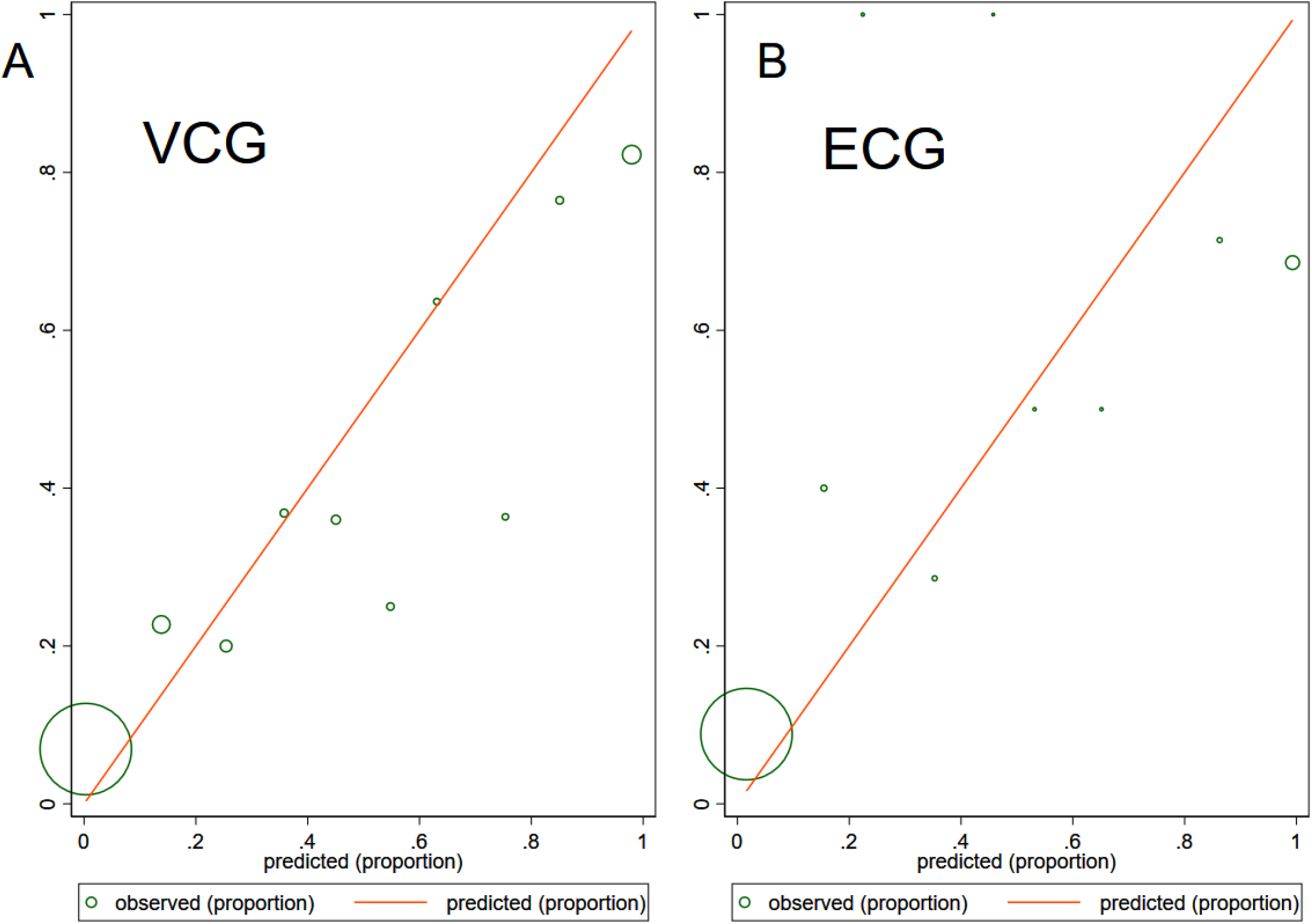
The calibration plot shows the observed and predicted CVD proportions in convolutional neural network model with (**A**) VCG input (43 variables) and (**B**) ECG input (153 variables). The size of the circles is proportional to the amount of data.

**Figure 14.**
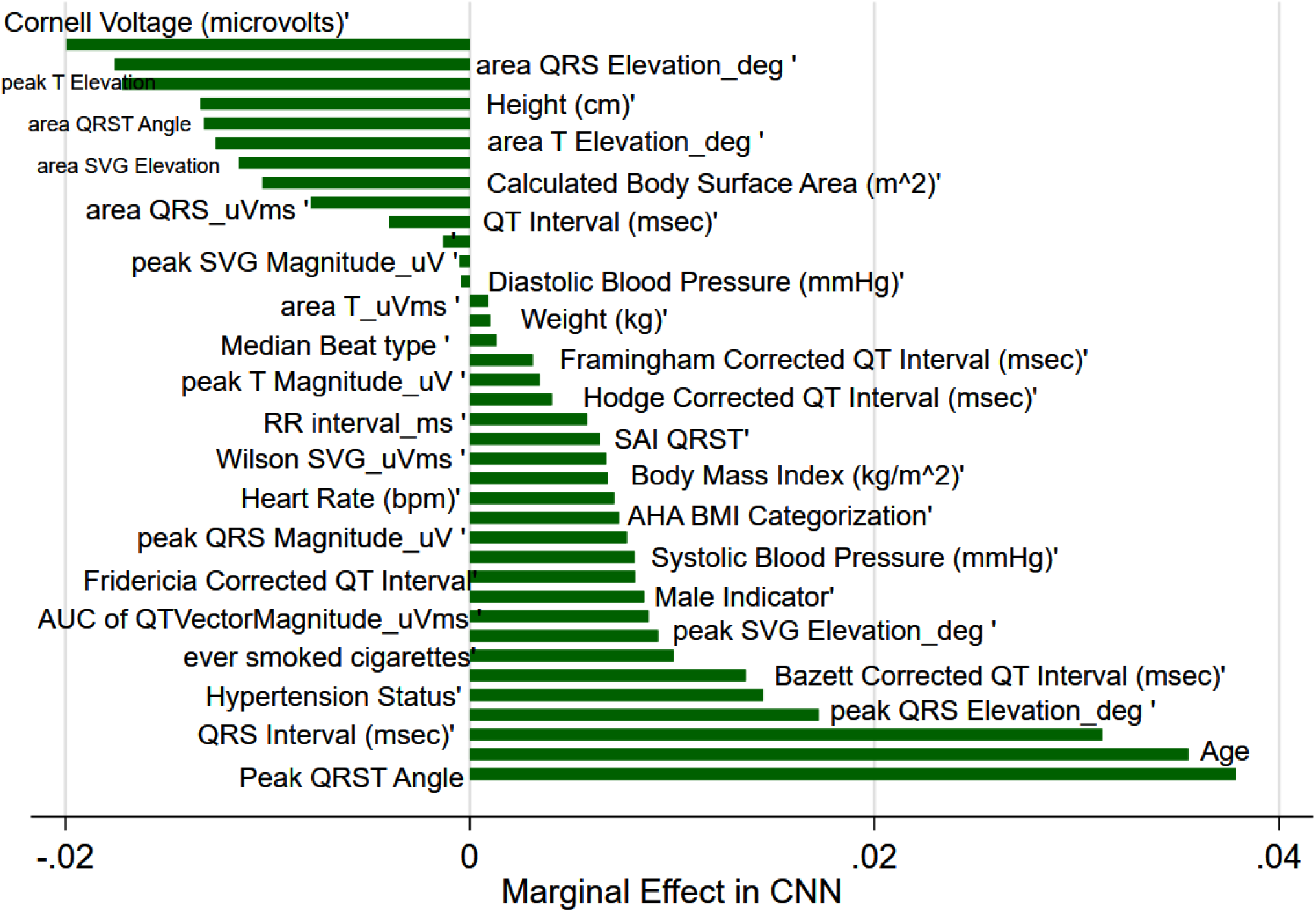
Comparison of the marginal effect size in a convolutional neural netrork with VCG input.

Several models (lasso, adaptive lasso, elastic net, ridge, and logistic regression) demonstrated an intermediate accuracy, similar fit and no differences in ROC AUC. Figure 15 shows cross-validation function and selected λ for each model. Selected predictors and their coefficients for all models are reported in Supplemental Table 3. Remarkably, the plug-in-based lasso model selected only two predictors: age and spatial peak QRS-T angle (Figure 16), while demonstrating only slightly smaller ROC AUC. The threshold of predictive function ≥ 0.026 identified all participants with prevalent CVD in the testing sample (100% sensitivity). Calibration of logistic regression (Figure 17), lasso (Figure 18), adaptive lasso (Figure 19), plug-in-based lasso (Figure 20), elastic net (Figure 21), and ridge (Figure 22) models was satisfactory.

**Figure 15.**
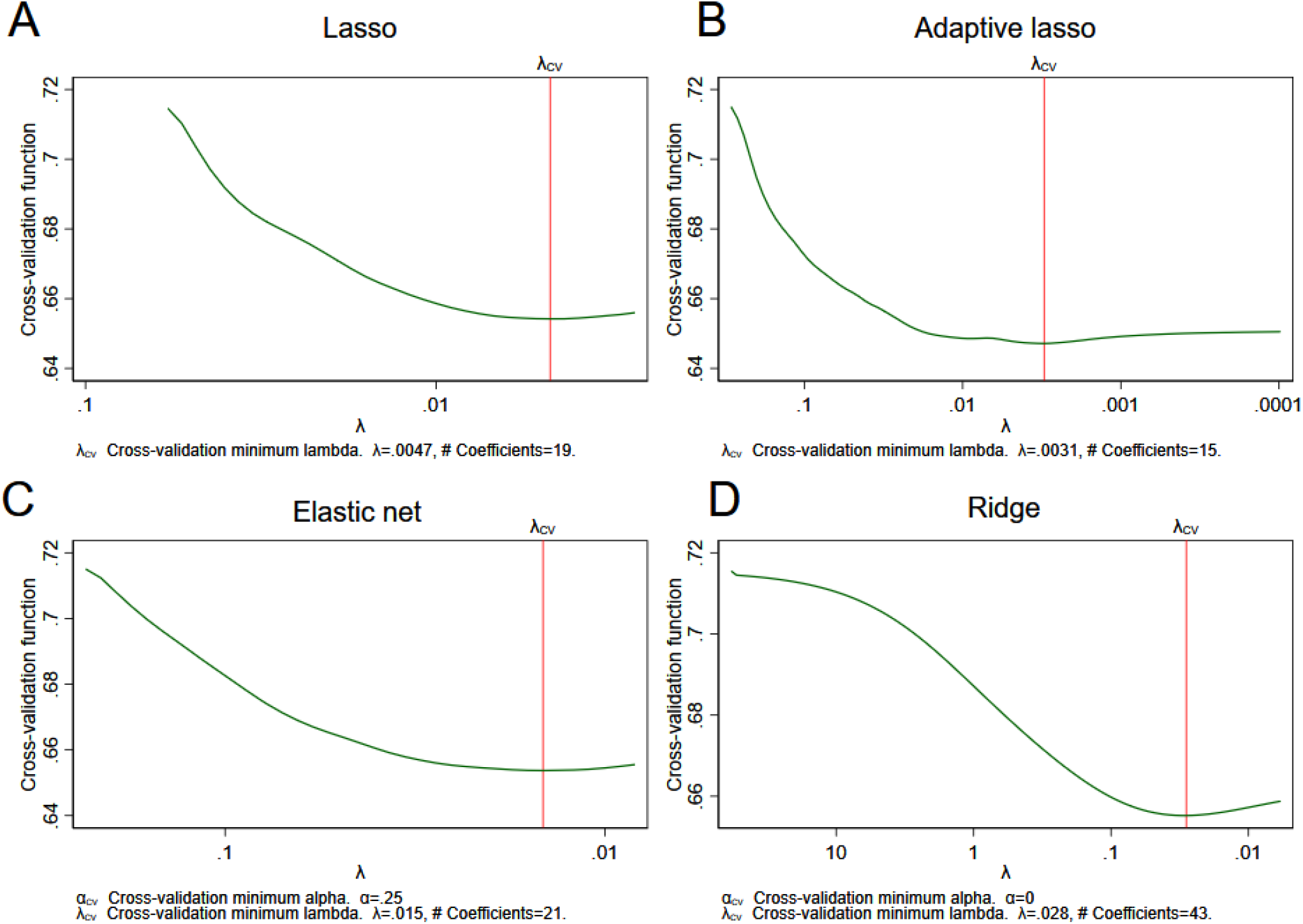
Cross-validation (CV) function (the mean deviance in the CV samples) is plotted over the search grid for the lasso penalty parameter λ on a reverse logarithmic scale for (**A**) lasso, (**B**) adaptive lasso, (**C**) elastic net, (**D**) ridge models. The first λ tried is on the left, and the last λ tried is on the right.

**Figure 16.**
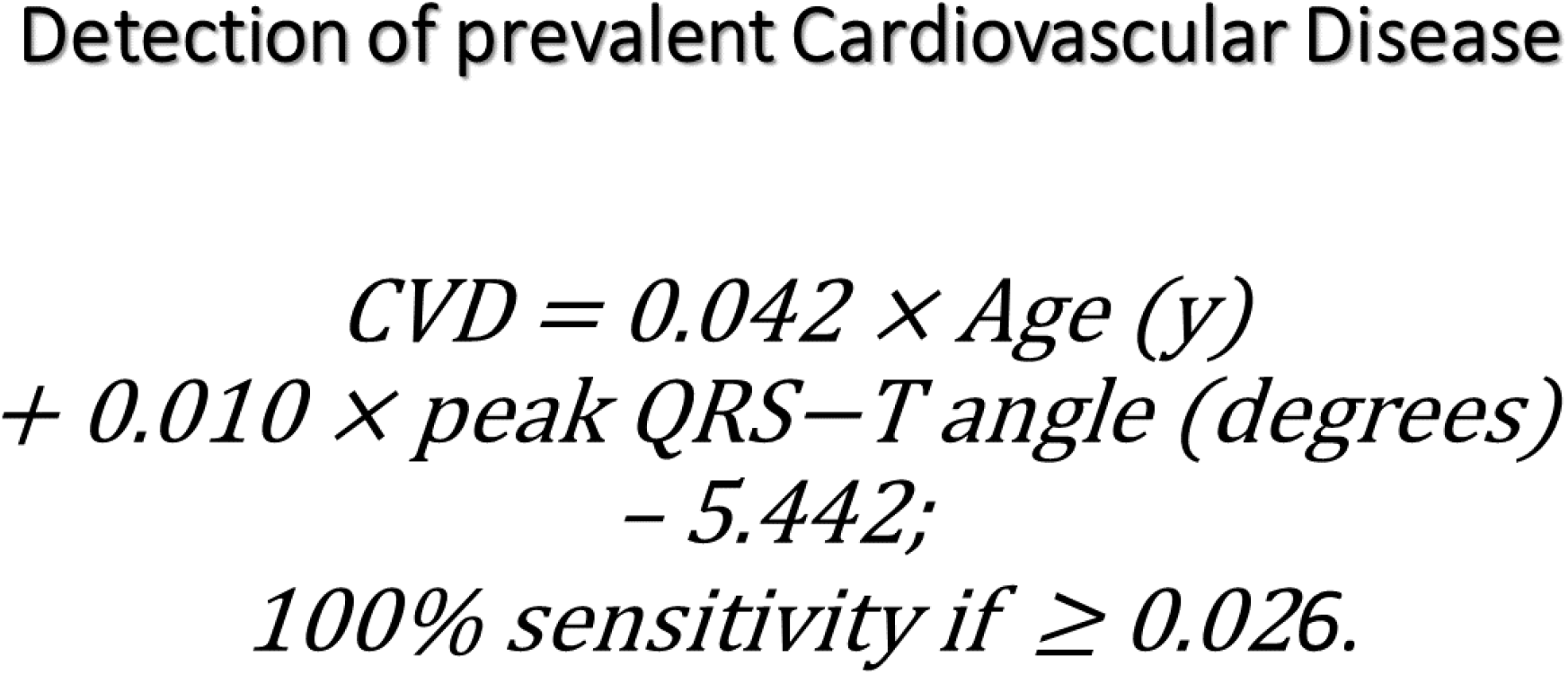
The final equation for the detection of prevalent CVD.

**Figure 17.**
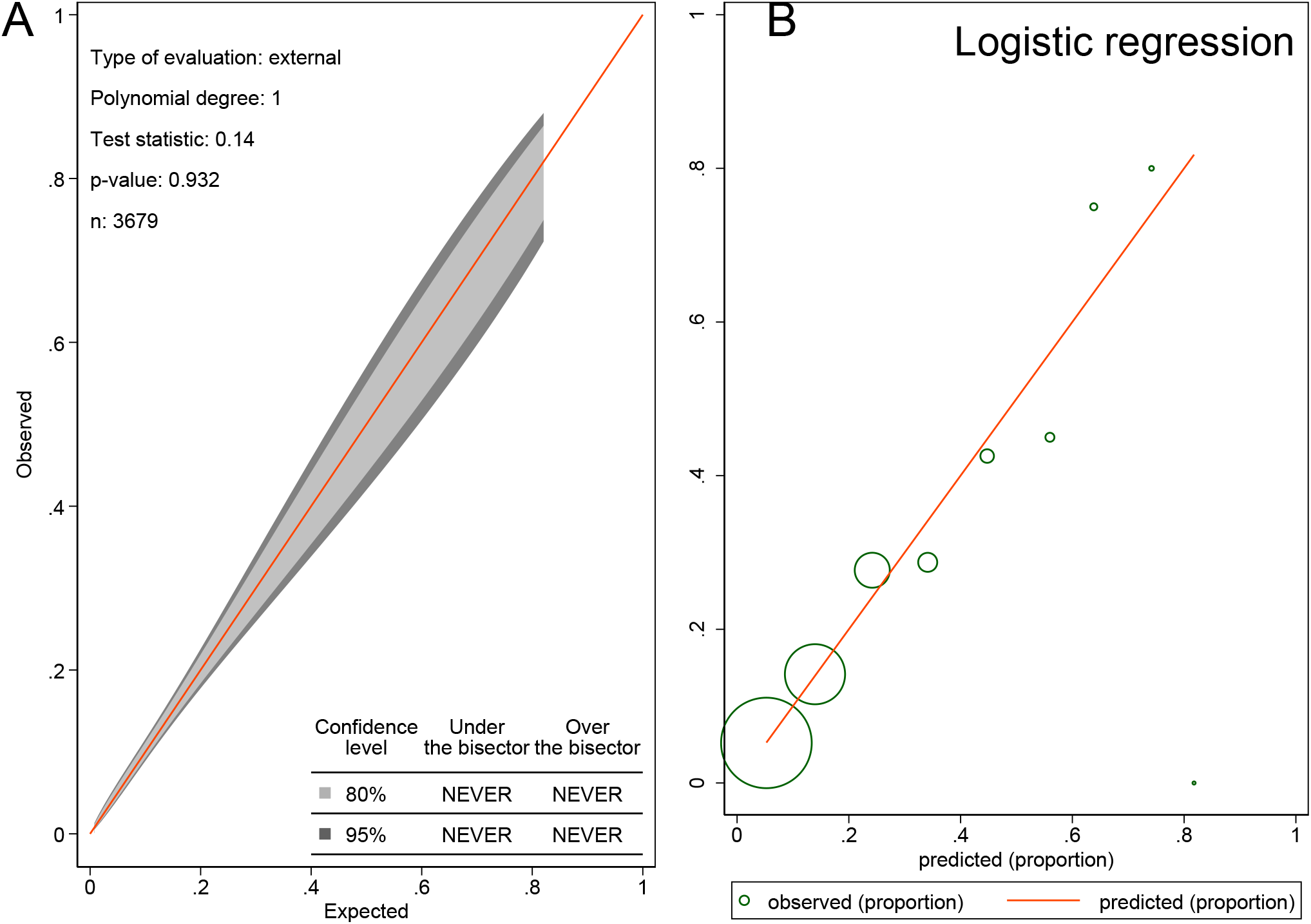
The calibration belt with 80% and 95% confidence intervals on the external sample (A) and the calibration plot (B) shows the observed and predicted CVD proportions in logistic regression model. The size of the circles is proportional to the amount of data.

**Figure 18.**
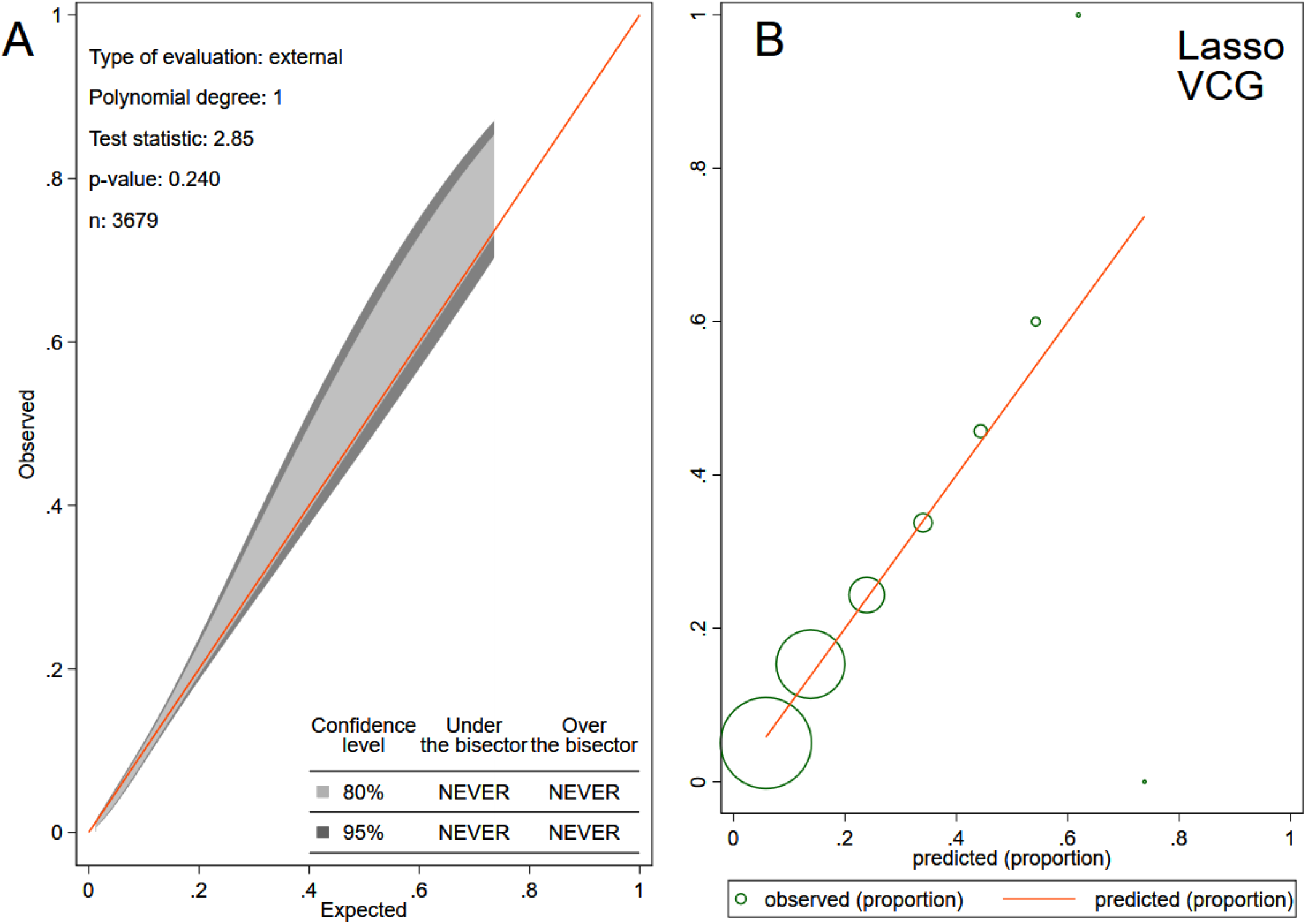
The calibration belt (A) and the calibration plot (B) shows the observed and predicted CVD in lasso model with VCG input. See Figure 17 legend for the details.

**Figure 19.**
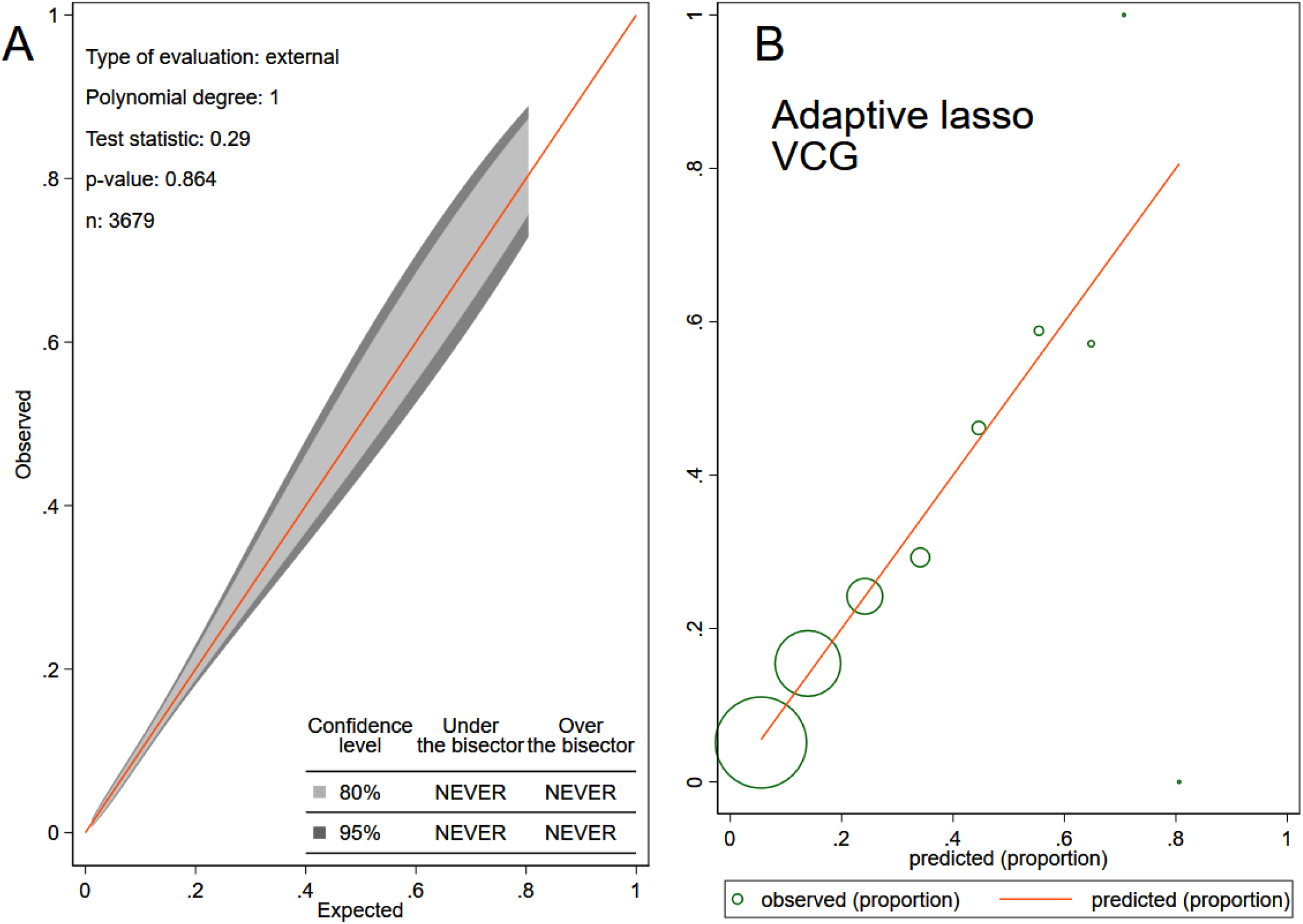
The calibration belt (A) and the calibration plot (B) shows the observed and predicted CVD in adaptive lasso model with VCG input. See Figure 17 legend for the details.

**Figure 20.**
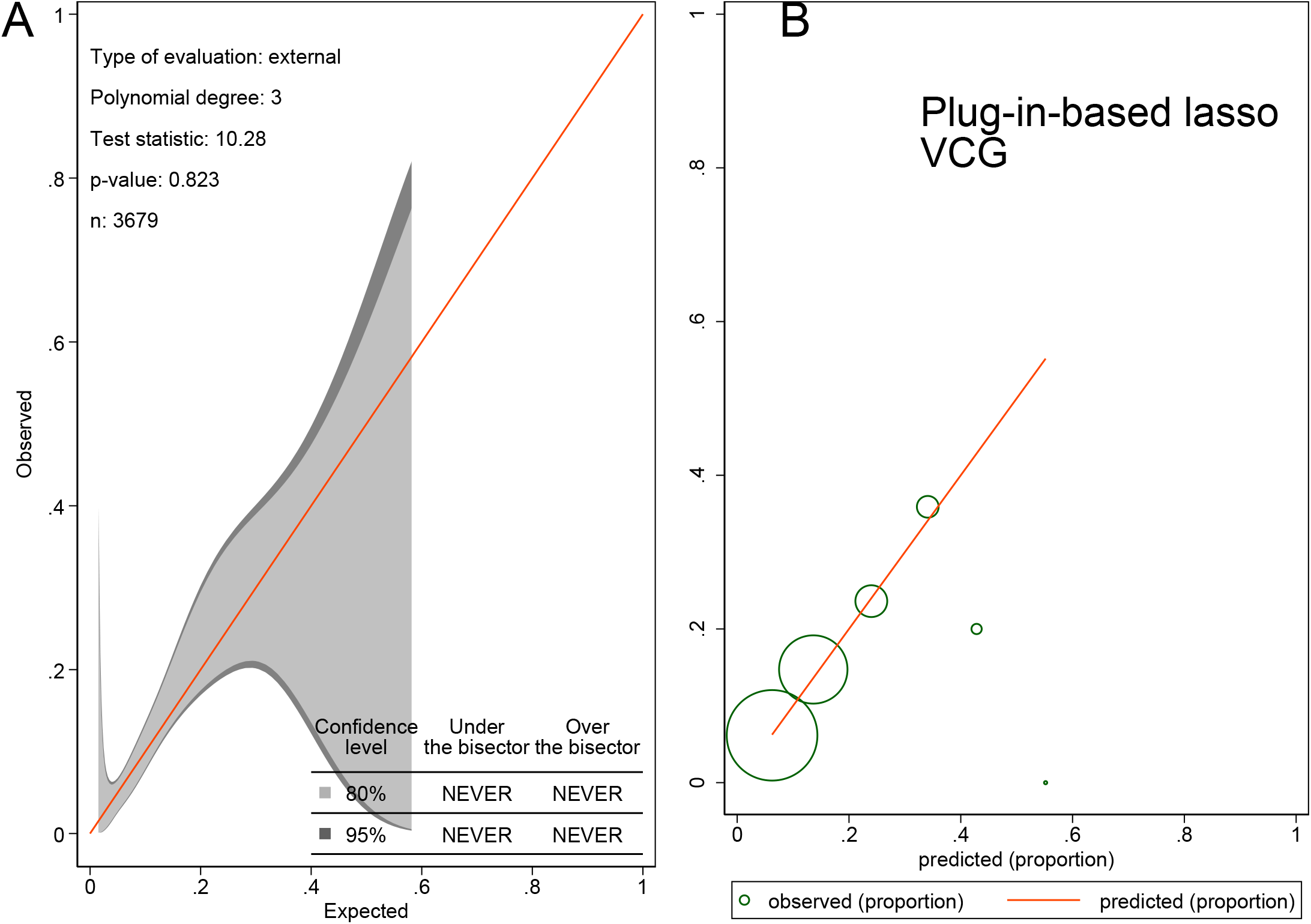
The calibration belt (A) and the calibration plot (B) shows the observed and predicted CVD in plug-in-based lasso model with VCG input. See Figure 17 legend for the details.

**Figure 21.**
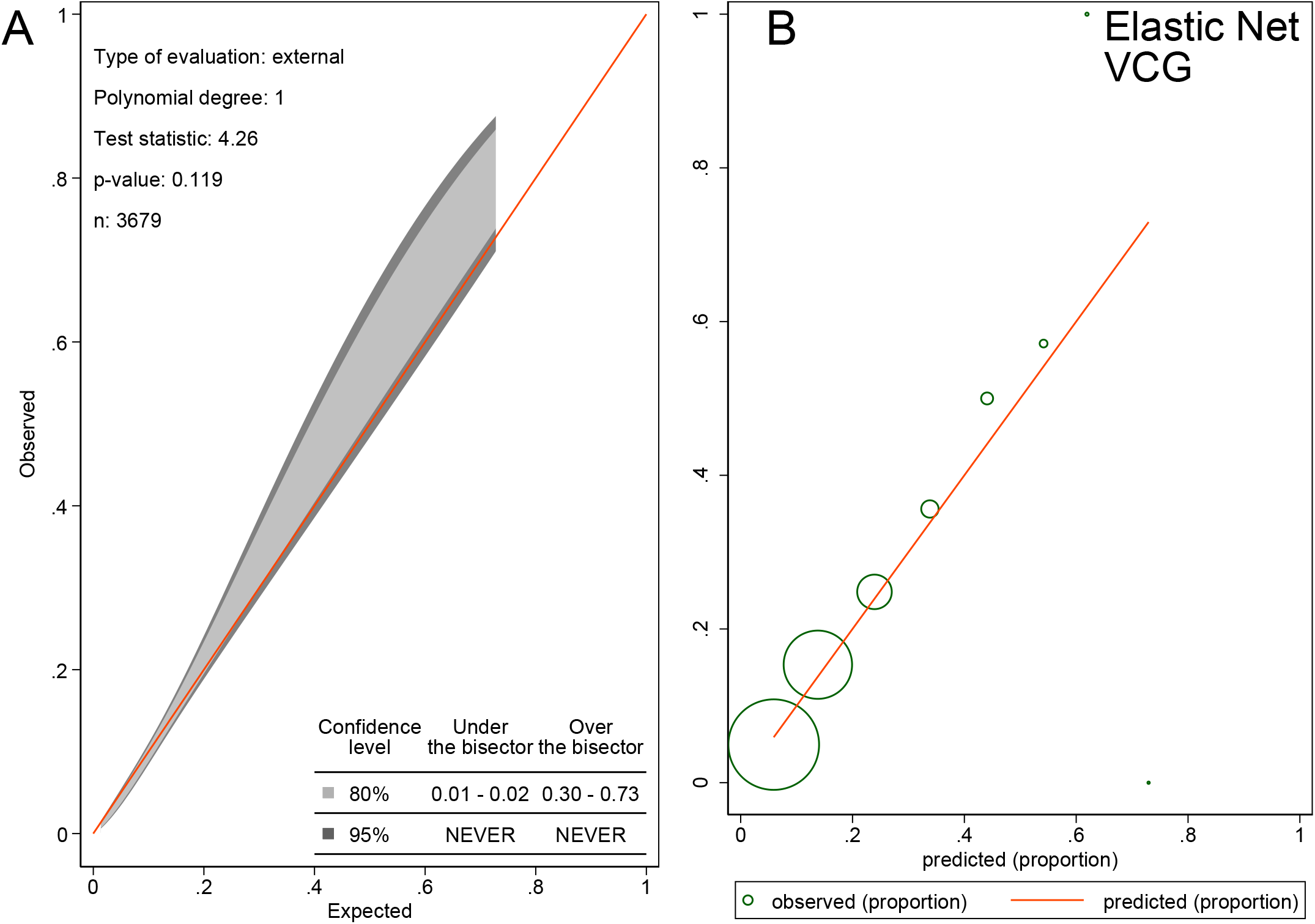
The calibration belt (A) and the calibration plot (B) shows the observed and predicted CVD in elastic net model with VCG input. See Figure 17 legend for the details.

**Figure 22.**
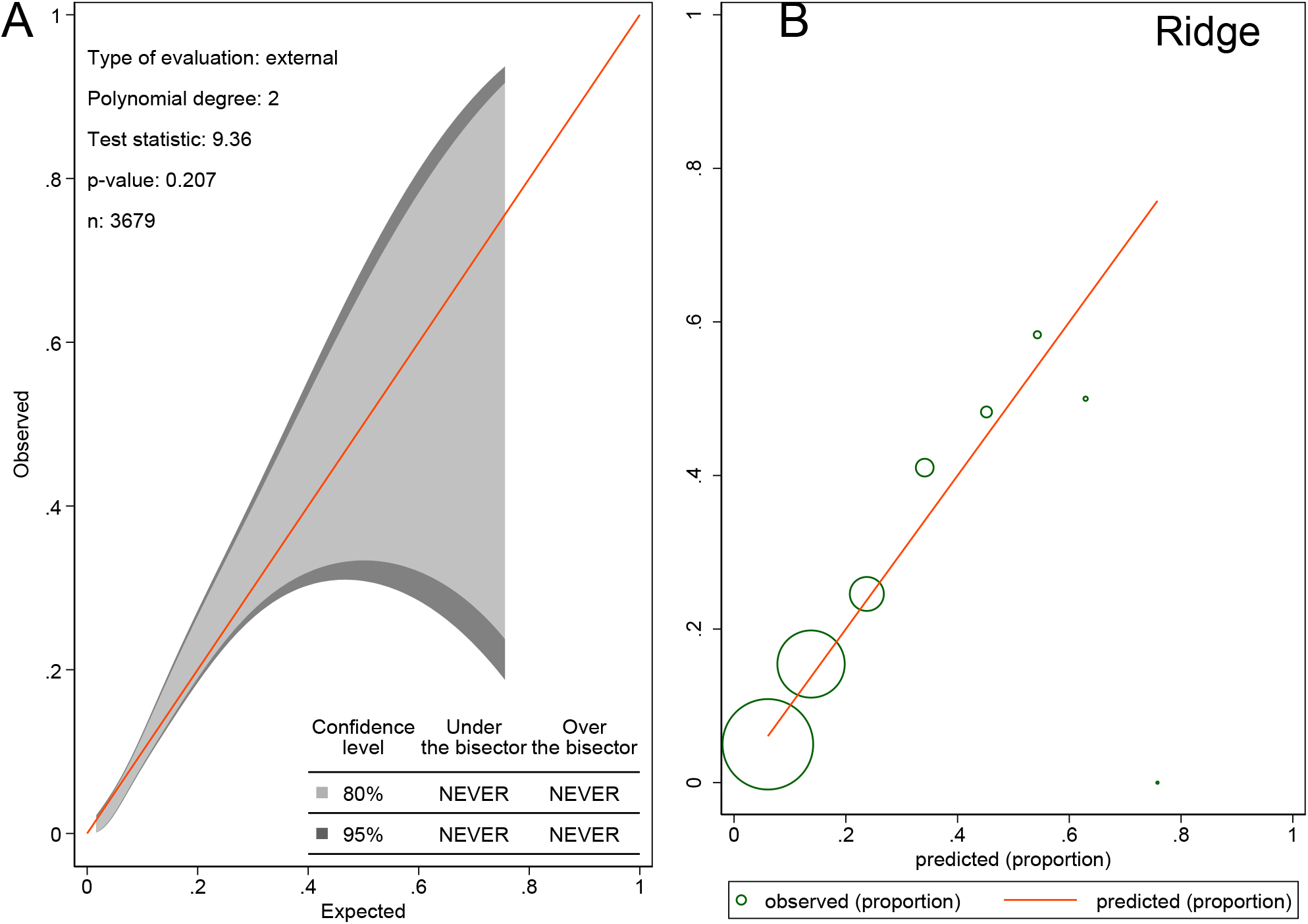
The calibration belt (A) and the calibration plot (B) shows the observed and predicted CVD in ridge model with VCG input. See Figure 17 legend for the details.

In validation out-of-sample population (Table 3), several models (logistic regression, lasso, adaptive lasso, elastic net, and ridge) had similarly high predictive accuracy, whereas convolutional neural network and plug-in-based lasso demonstrated slightly, but statistically significantly lower accuracy. A pre-selected threshold of plug-in-based lasso predictive function was 100% sensitive and identified all participants with prevalent CVD in the validation sample. Random forests model performance was unsatisfactory.

### Comparison of machine learning models with the input of VCG and 12-lead ECG features

Selected predictor variables and beta-coefficients were reported in Supplemental Table 4. Lasso family models selected 5-79 predictors, which included finicky features of ECG (P-prime, Q, and R-prime measurements). In a training and testing sample, all models that included both VCG and ECG predictors showed higher accuracy than VCG-only models (Table 3). However, there was no difference in ROC AUC between respective models in validation sample.

Furthermore, only plug-in-based lasso and adaptive lasso models showed satisfactory calibration (Figures 23-24), whereas calibration of elastic net and lasso models became unsatisfactory (Figure 25-26).

**Figure 23.**
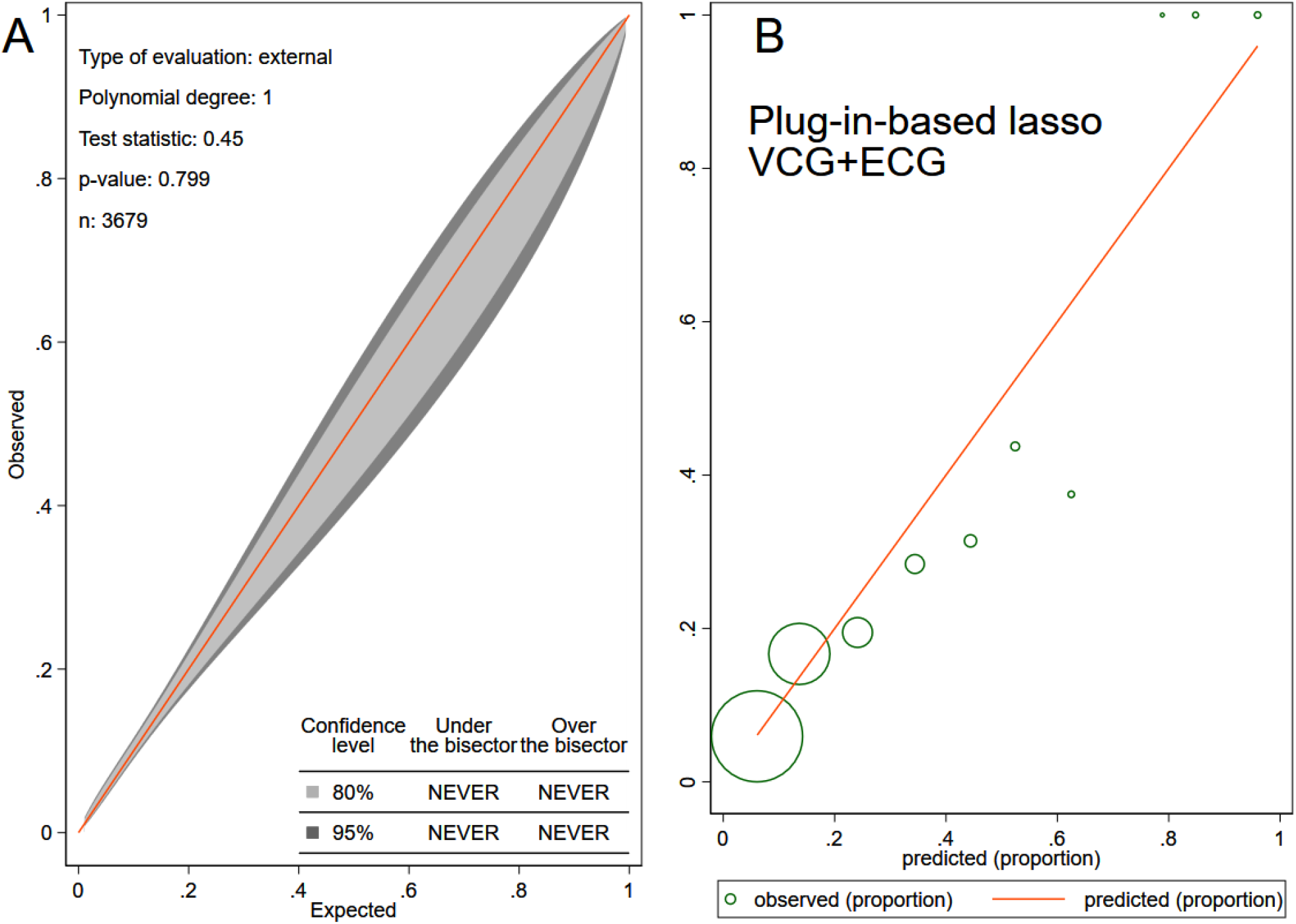
The calibration belt (A) and the calibration plot (B) shows the observed and predicted CVD in plug-in-based lasso model with VCG and ECG input. See Figure 17 legend for the details.

**Figure 24.**
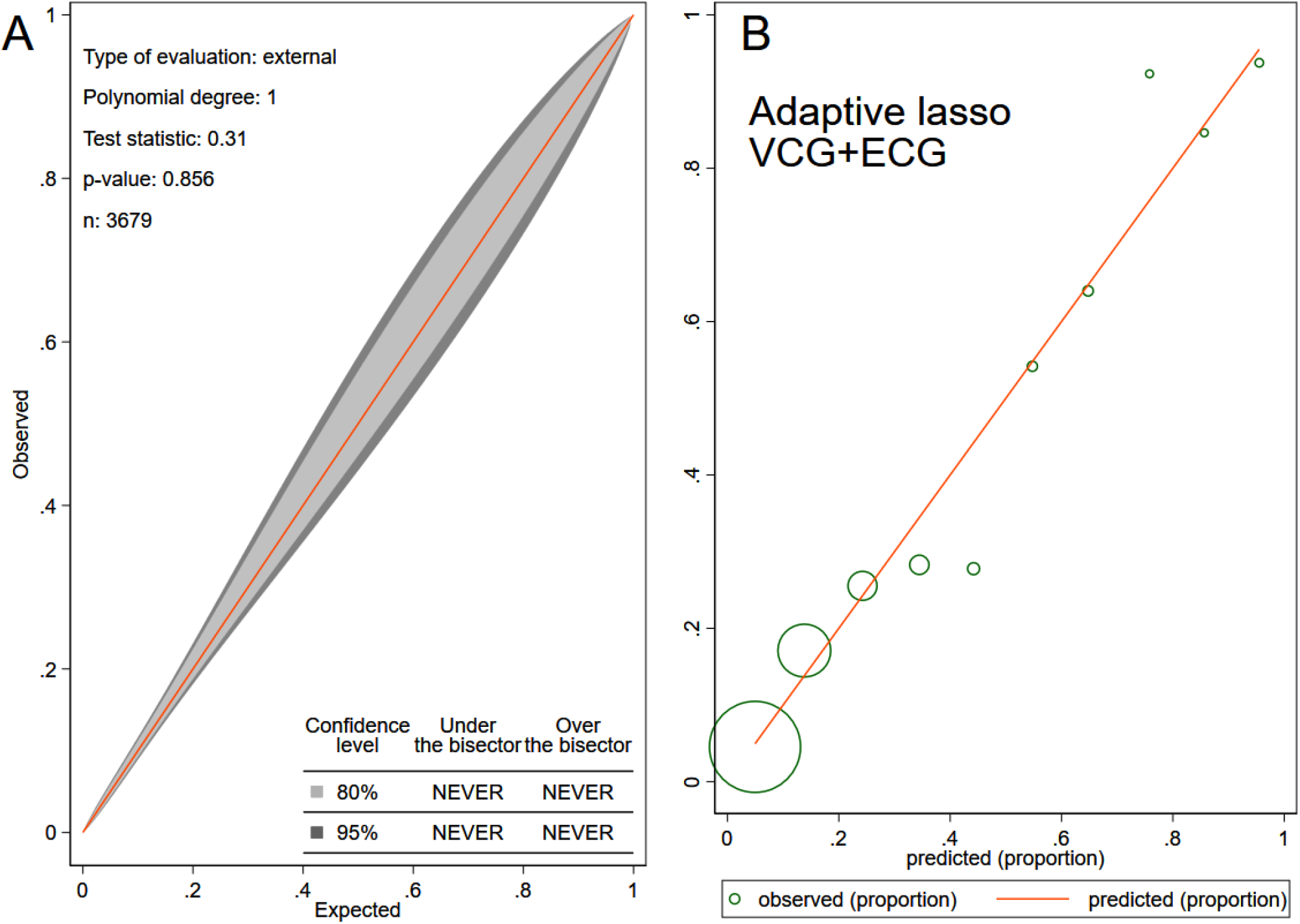
The calibration belt (A) and the calibration plot (B) shows the observed and predicted CVD in adaptive lasso model with VCG and ECG input. See Figure 17 legend for the details.

**Figure 25.**
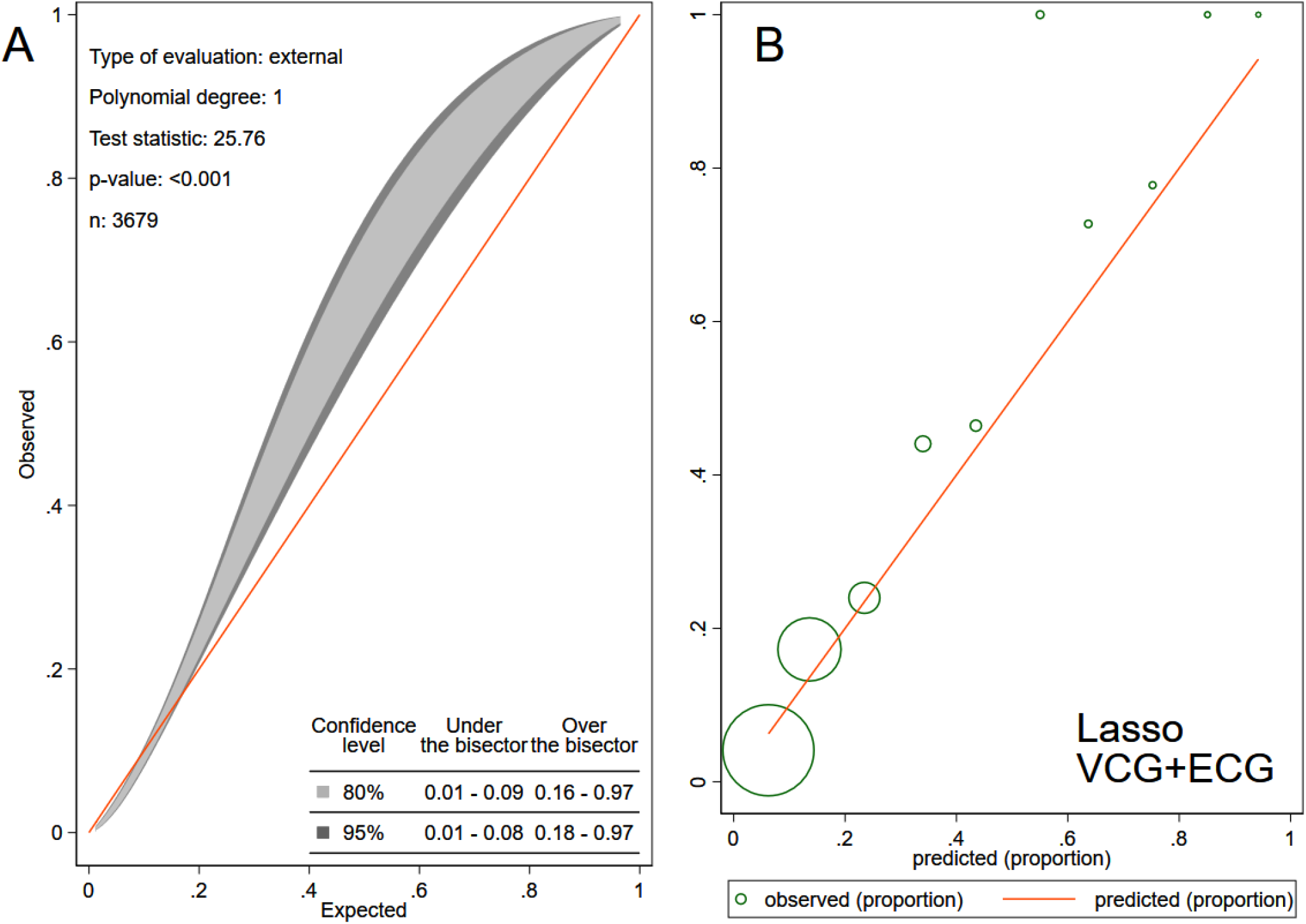
The calibration belt (A) and the calibration plot (B) shows the observed and predicted CVD in lasso model with VCG and ECG input. See Figure 17 legend for the details.

**Figure 26.**
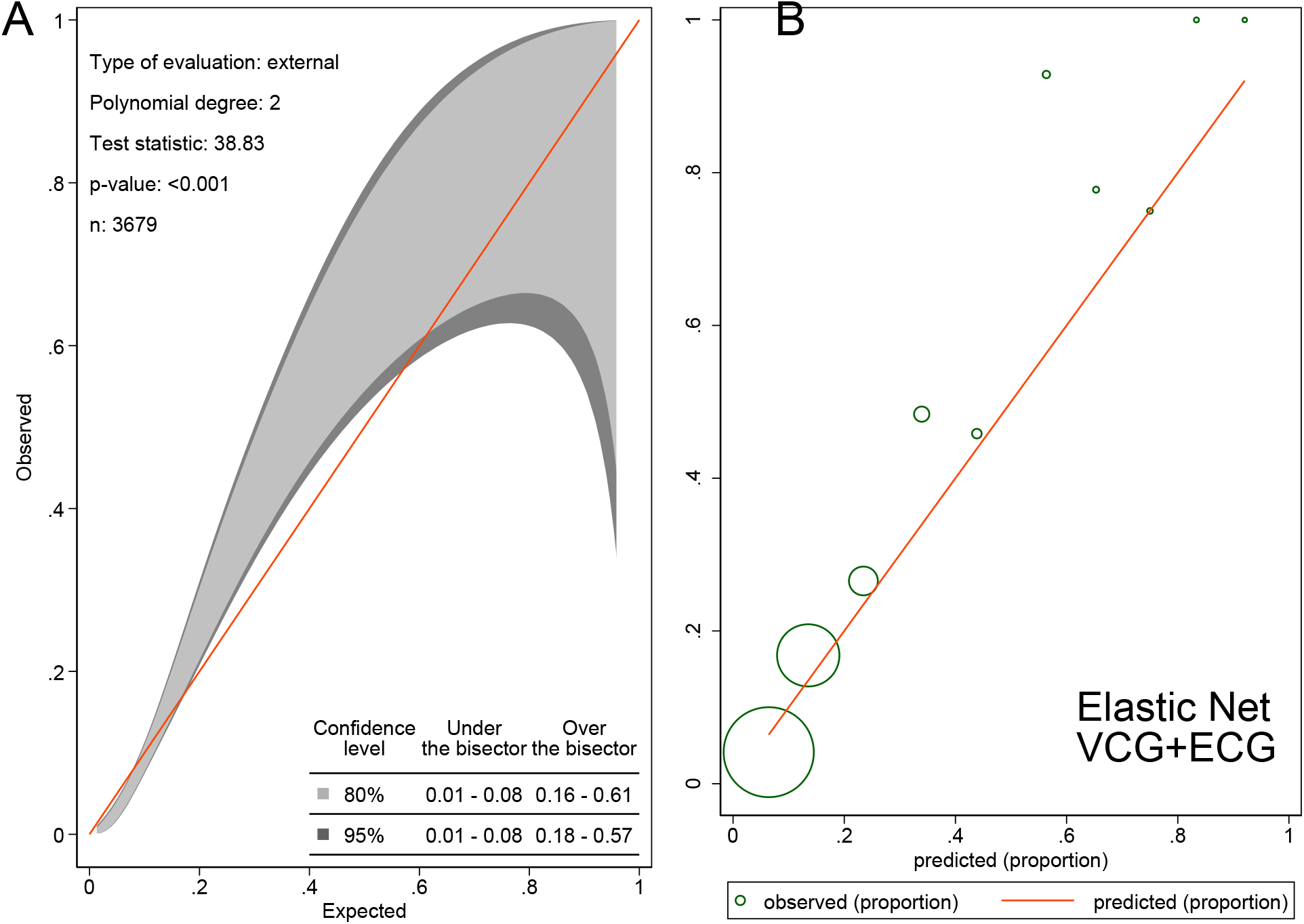
The calibration belt (A) and the calibration plot (B) shows the observed and predicted CVD in elastic net model with VCG and ECG input. See Figure 17 legend for the details.

Random forest model with 695 input variables that included both ECG and VCG predictors was tuned (Figure 27) and included 500 subtrees and 26 variables to randomly investigate at each split. In validation sample, the final VCG+ECG random forest model reported smaller error (10%) than VCG-based model. The model correctly detected CVD in only 14 out of 75 individuals (sensitivity 19%), while it accurately identified all 536 CVD-free participants (specificity 100%). The most influential predictors are shown in Figure 28.

**Figure 27.**
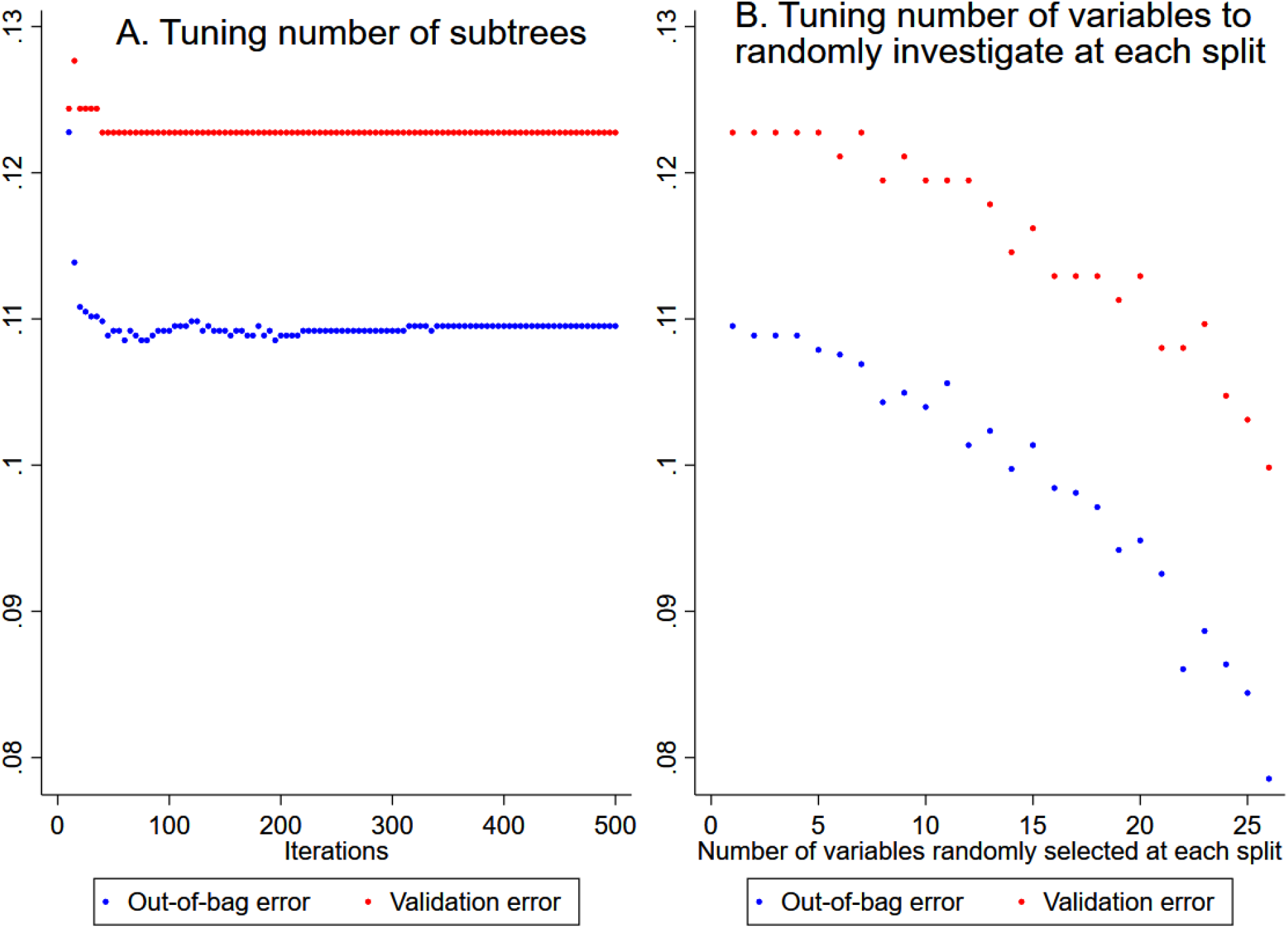
Out-of-bag error and validation error plotted versus (**A**) number of iterations or subtrees, and (**B**) number of variables randomly investigated at each split in a random forest model with both VCG and ECG input (695 variables).

**Figure 28.**
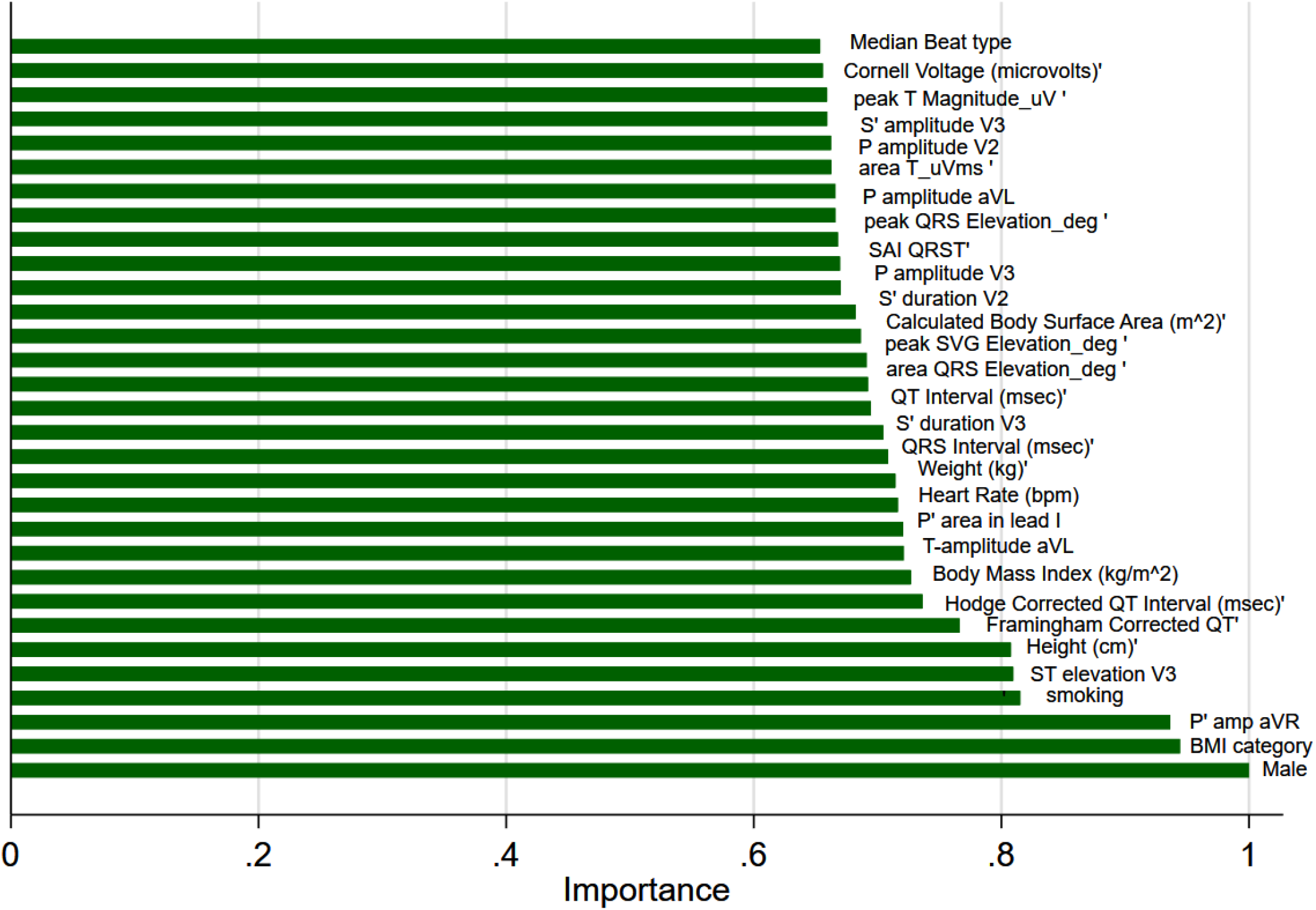
Importance scores of the most important predictor variables in a random forest model with both VCG and ECG input.

The convolutional neural network with the input of 153 ECG predictor variables demonstrated moderate predictive accuracy, which was significantly worse as compared to the convolutional neural network model with VCG input (Table 3), and poor calibration (Figure 13 B).

## Discussion

This large community-based cross-sectional study of nearly 4,000 African American men and women revealed several novel findings. First, we demonstrated sex differences in GEH after rigorous adjustment for prevalent CVD, cardiovascular risk factors, sociodemographic, and anthropometric characteristics. Secondly, we showed that sex modified an association of CVD with ECG and VCG phenotype. Thirdly, we observed a significant effect of unmeasured genetic and environmental factors on T and SVG azimuth. The azimuth of T and SVG vectors can serve as sensitive markers of cardiac repolarization. Finally, we developed and validated a simple model for the detection of prevalent CVD, which included age and QRS-T angle. In the future, automated ECG measurements could be implemented in barbershops. Our findings open an avenue for the development of pharmacist-led interventions for secondary prevention of CVD in barbershops and may ultimately reduce cardiovascular morbidity and mortality in AA communities.

### Sex differences in GEH

Consistently with the recent study in the ARIC cohort^12^, we observed significant sex differences in the SAI QRST and the SVG vector direction, but not in peak SVG magnitude. Importantly, the size of sex differences previously seen in the predominantly white populations^12, 23, 26^, was mainly similar to that found in AA men and women in this study. Sex is biologically determined, and sex differences in GEH manifest independently of race.^11^ In this study, we showed sex differences in GEH that persisted after adjustment for anthropometric characteristics, prevalent CVD, and cardiovascular risk factors, suggesting that GEH can detect sex differences in the underlying expression of potassium channels.^45, 46^ Consistently with our findings, recent analysis of the double-blind placebo-controlled trial showed that dofetilide had a larger effect on the spatial QRS-T angle in women than in men.^47^

### Sex modifies an association of CVD with ECG and VCG phenotype

Our study corroborated a well-known association of GEH with CVD.^28, 29, 37, 38^ However, little was known about how sex modifies an association of GEH with CVD. In a prospective cohort study conducted in the predominantly white Finnish population, SAI QRST strongly associated with cardiovascular mortality in women, but not in men.^16^ In accordance with Lipponen et al^16^, our cross-sectional study observed differences in SAI QRST by CVD status in women, but not in men. In ARIC, the spatial QRS-T angle was more strongly associated with fatal CHD in women than in men.^48^ In contrast, sex did not modify an association of QRS-T angle with prevalent CVD in this study, which can be explained by differences in the definition of outcome. In the bi-racial ARIC population, a substantial number of ECG markers (QRS duration, Cornell voltage, SAI QRST, SVG magnitude, heart rate, and QTc) were associated with a larger risk of SCD in women than in men.^12^ Notably, we newly observed smaller Wilson’s SVG, T area, and T peak magnitude in men with CVD as compared to CVD-free men, but no differences in women. Altogether, our study showed that sex significantly modifies an association of prevalent CVD with GEH.

### An effect of unmeasured genetic and environmental factors on repolarization

Our study, for the first time, showed significant random effects carried by family units, manifested by a substantial range of differences in T and SVG azimuth between families. Some family units had very large differences (up to 40 degrees) in T loop direction between family members with different characteristics (e.g. male vs. female; with vs. without CVD), whereas other family units either had very little differences in T loop direction between family members with different characteristics, or those differences were in an opposite direction. This study cannot answer whether observed differences were due to underlying genetic variations or different environmental exposures. Numerous pharmacological, dietary^49, 50^, and environmental factors can block the cardiac human ether-à-go-go-related gene (HERG) channel^51, 52^, which can explain differences in repolarization characteristics between families. On the other hand, a previous JHS study^53^ showed a common genetic variant *SCN5A-1103Y* was associated with prolongation of the QT interval and shortening of QRS. In the JHS, 15% of AA participants are carriers of *SCN5A-1103Y*.^53^ Intriguingly, a mixed model with random intercept was the optimal fit for QRS duration in this study, suggesting the importance of between-families differences in QRS. Further studies of the effects of environmental exposures and genetic variations on GEH in the JHS are needed.

### Automated detection of prevalent CVD using machine learning

In this study, we used ML to detect prevalent CVD (Figure 29). We developed and validated a model that consists of readily available parameters – age and QRS-T angle – which robustly detected prevalent CVD with an accuracy of approximately 0.7. Our finding opens an avenue for a randomized controlled trial of pharmacist-led interventions (e.g., statins, aspirin, BP-lowering drugs) in barbershops, for secondary prevention of CVD in AA communities.

**Figure 29.**
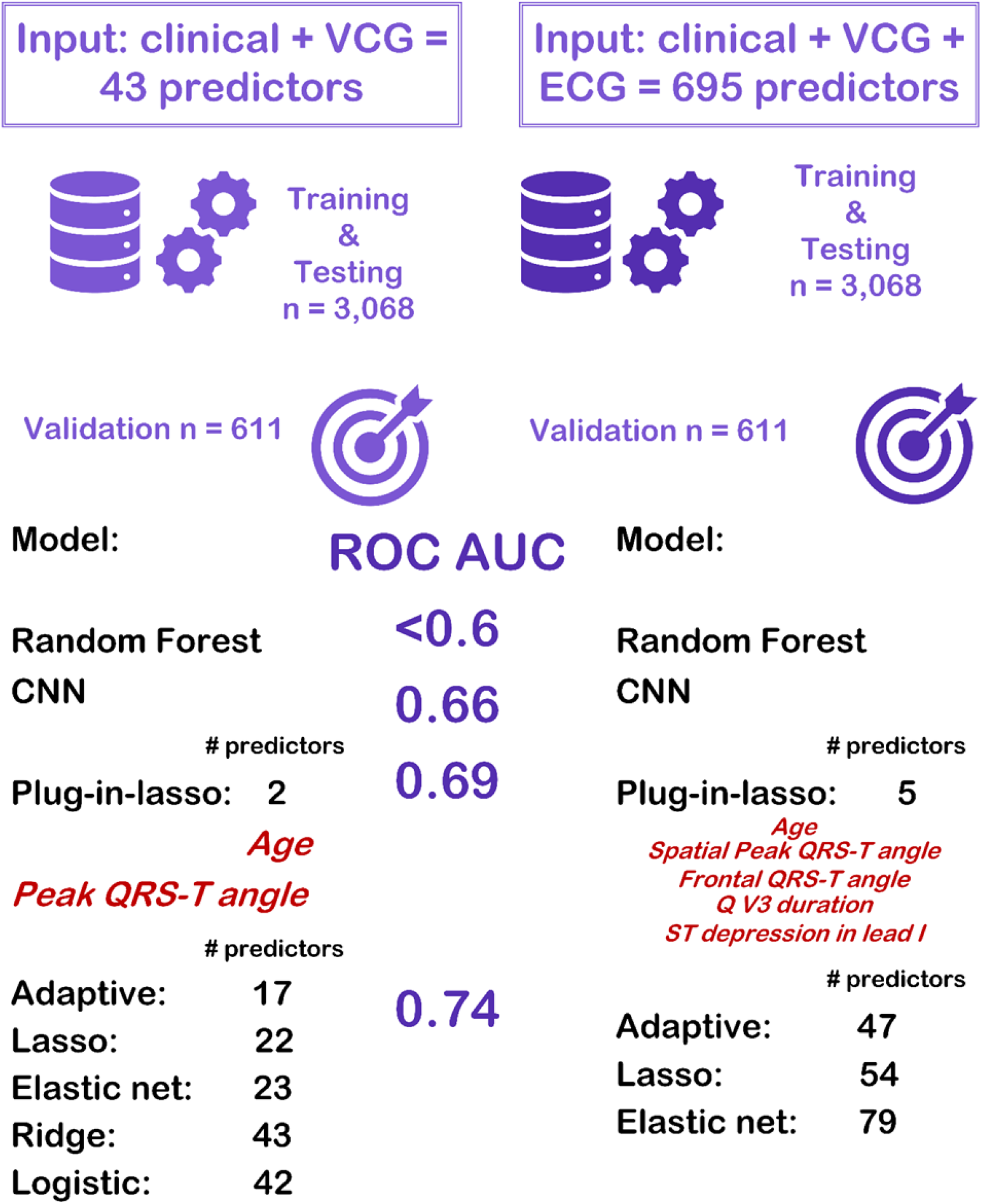
Schematic presentation of machine learning results

Overwhelming data have proved that the use of statins, aspirin, and BP-lowering medications for secondary prevention of CVD reduces mortality.^54^ However, in the United States, among CVD patients, only 45% receive aspirin, 88% receive antihypertensive medication, and 65% receive statins.^55^ Furthermore, adherence to statin use is low, especially in AA adults.^56^ Among AAs, CVD is underdiagnosed and undertreated.^2^ Screening for prevalent CVD in barbershops with subsequent pharmacist-led interventions can save thousands of lives in AA communities. A randomized clinical trial is warranted to test the proposed strategy.

In this study, the ML approach selected QRS-T angle as the most important predictor, which, together with age, is both necessary and sufficient for the detection of prevalent CVD. It is remarkable that the QRS-T angle outperformed well-known CVD risk markers, including hypertension, smoking, and BMI, which highlights the importance of information carried by VCG. Interestingly, Jensen et al.^11^ showed that the spatial QRS-T angle was the only GEH parameter that interacted with race in the association with SCD.^11^ After rigorous adjustment for CVD and cardiovascular risk factors, the QRS-T angle was associated with SCD in white, but not in black ARIC study participants.^11^ This finding is consistent with our results, showing the strongest association of spatial QRS-T angle with prevalent CVD in AA men and women.

The Personalized Risk Identification and Management for Arrhythmias and Heart Failure by ECG and CMR (PRIMERI) study^57^ prospectively enrolled participants (40% AAs) with spatial QRS-T angle ≥ 105°or Selvester score ≥ 5 and showed that more than half of them had myocardial scar.^58^

Importantly, our study compared the performance of models selected by supervised ML with two sets of input variables. We found that the models using the input of nearly 700 ECG features selected finicky, rarely observed ECG features (e.g., P-prime in V2-V5, R-prime in lead I and aVR), and did not improve final VCG-based models, which selected global VCG features that describe the directions of QRS, T, and SVG vectors. For all models, the set of input variables determined the final selection of variables.

While the ML approach is gaining strengths in cardiology^59-61^, only one previous study used ML for the detection of prevalent CVD. Dinh et al.^62^ used an input of 131 clinical characteristics in the National Health and Nutrition Examination Survey (NHANES) data and reported ROC AUC of ∼ 0.8. Unfortunately, Dinh et al.^62^ did not report β-coefficients for the selected final 24 features, which made impossible external validation of their findings. Also, many of the selected NHANES features are prone to recall bias (e.g. dietary habits: carbohydrate, calcium, fiber, caffeine, sodium intake), and are burdensome for participants.

### Strengths and Limitations

The strengths of the study include its design of a large community study of AA adults with the nested family cohort as well as well-validated definitions of prevalent CVD and traditional cardiovascular risk factors. However, the study limitations have to be acknowledged. We conducted a cross-sectional analysis, which precluded the causal interpretation of the observed associations. As we did not validate the relatedness of the study participants comprising family units, we were not able to separate the effects of genetic and environmental factors. It is possible that unrelated people were included in a family unit by a random chance. Excluded from the regression analysis participants with missing covariates were somewhat different from those who were included. Nevertheless, the study obtained meaningful results, using appropriate analytical approaches.

## Conclusions and Clinical Implications

In summary, in this study, we showed that the prevalent CVD is associated with GEH after adjustment for demographic, anthropometric, socioeconomic, and traditional cardiovascular risk factors, and sex modifies an association of prevalent CVD with GEH. Our study provided new evidence of sex differences in the electrical signature of CVD, reflecting unique underlying biological pathways in AA men and women with and without CVD. VCG and GEH characteristics added multidimensionality in the description of the sex differences. When compared with men, women’s SVG points farther posteriorly and more downward. Women with CVD have larger SAI QRST than CVD-free women. In contrast, men with CVD have smaller Tarea than CVD-free men. Importantly, we described a range of differences in the direction of the cardiac repolarization vector in response to unmeasured environmental exposures and genetic variations. Observed sex differences support sex-specific approaches to CVD prediction, prevention, and management in AA men and women.

Furthermore, in this study, we developed and validated a simple model for CVD detection, comprised of age and QRS-T angle. There is a 70% chance that our model is able to distinguish between CVD presence or absence. We selected a cut-off that corresponds to 100% sensitivity, to make it more useful for screening. In the future, our test can be used in barbershops, churches, and other community centers. A strategy of providing availability for CVD detection in AA communities with subsequent pharmacist-led interventions for secondary prevention of CVD should be tested in future clinical trials.

## Data Availability

The JHS data are available through the National Heart, Lung, and Blood Institute Biological Specimen and Data Repository Information Coordinating Center (BioLINCC) and the National Center of Biotechnology Information database of Genotypes and Phenotypes (dbGaP).

## Acknowledgment

The authors thank the staff and participants of the JHS. We thank Francis Phan, MD, and John Johnson, BS, for their help with ECG analyses.

---

**Supplemental Table 1.** Comparison of included vs. excluded participants

| Characteristics | Included (n=3,001) | Excluded (n=678) | P-value |
| --- | --- | --- | --- |
| Age(SD), y | 62.4(11.5) | 58.3(13.2) | <0.0001 |
| Male, n(%) | 1,151(38.4) | 181(26.7) | <0.0001 |
| Weight(SD), kg | 91.1(21.3) | 92.0(22.4) | 0.302 |
| Height(SD), cm | 168.8(9.5) | 167.2(9.0) | 0.0001 |
| BMI(SD), kg/m<sup>2</sup> | 32.0(7.0) | 33.0(8.0) | 0.002 |
| BSA(SD), m<sup>2</sup> | 2.00(0.24) | 2.00(0.24) | 0.529 |
| Ever tobacco smoker, n(%) | 906(30.2) | 191(28.2) | 0.299 |
| Hypertension, n(%) | 2,225(74.1) | 478(70.5) | 0.052 |
| Systolic blood pressure(SD), mmHg | 127.9(18.5) | 126.2(19.5) | 0.039 |
| Diastolic blood pressure (SD), mmHg | 75.2(10.7) | 74.5(11.6) | 0.139 |
| Heart rate(SD), bpm | 63.6(10.5) | 65.5(11.5) | 0.0001 |
| QRS duration(SD), ms | 89.2(15.7) | 88.2(15.3) | 0.126 |
| Bazett corrected QT (SD), ms | 426.5(25.5) | 427.9(26.7) | 0.191 |
| Cornell voltage(SD), mV | 1.53(0.59) | 1.43(0.61) | <0.0001 |
| Median beat: normal sinus, n(%) | 2,957(98.5) | 672(99.1) |  |
| Median beat: atrial fibrillation, n(%) | 12(0.4) | 2(0.3) | 0.482 |
| Median beat: ventricular pacing, n(%) | 32(1.1) | 4(0.6) |  |
| QRS area(SD), mV*ms | 38.5(18.3) | 38.1(18.2) | 0.573 |
| Peak QRS magnitude(SD), mV | 1.59(0.43) | 1.57(0.45) | 0.293 |
| Area QRS elevation(95%CI),° | 73.8(73.2-74.4) | 71.3(69.9-72.7) | <0.001 |
| Peak QRS elevation(95%CI),° | 72.7(72.2-73.2) | 70.4(69.3-71.6) | <0.001 |
| T area(SD), mV*ms | 48.7(23.4) | 47.6(22.0) | 0.270 |
| Peak T magnitude(SD), mV | 0.36(0.16) | 0.36(0.15) | 0.971 |
| Area SVG azimuth(95%CI),° | -14.3(-15.1 to -14.0) | -12.8(-14.7 to -11.0) | >0.05 |
| Peak SVG azimuth(95%CI),° | 3.4(2.6-4.1) | 5.0(3.3-6.7) | >0.05 |
| Wilson's (area) SVG (SD), mV*ms | 69.6(28.5) | 68.2(27.0) | 0.243 |
| Peak SVG magnitude (SD), mV | 1.81(0.50) | 1.80(0.50) | 0.433 |
| Area SVG elevation (95%CI),° | 71.8(71.3-72.3) | 70.4(69.2-71.6) | <0.01 |
| Peak SVG elevation (95%CI),° | 71.2(70.7-71.6) | 69.2(68.1-70.3) | <0.001 |
| SAI QRST(SD), mV*ms | 154.5(51.7) | 151.2(50.2) | 0.127 |
| Area QRS-T angle(SD),° | 69.3(35.5) | 68.4(36.6) | 0.540 |
| Peak QRS-T angle(SD),° | 52.5(38.4) | 52.5(39.4) | 0.999 |

**Supplemental Table 2.** Comparison of training & testing, and validation groups

| Characteristics | All (n=3,679) | Training (n=3,068) | Validation (n=611) | P-value |
| --- | --- | --- | --- | --- |
| Age(SD), y | 61.6(11.9) | 61.4(11.9) | 62.6(11.6) | 0.025 |
| Male, n(%) | 1,332(36.2) | 1,109(36.2) | 223(36.5) | 0.869 |
| Weight(SD), kg | 91.2(21.5) | 91.2(21.6) | 91.3(21.5) | 0.991 |
| Height(SD), cm | 168.5(9.4) | 168.5(9.4) | 168.6(9.3) | 0.786 |
| BMI(SD), kg/m<sup>2</sup> | 32.1(7.2) | 32.1(7.2) | 32.1(7.4) | 0.970 |
| Obese BMI group, n(%) | 2,084(56.7) | 1,738(56.6) | 346(56.6) | 0.966 |
| BSA(SD), m<sup>2</sup> | 2.00(0.25) | 2.00(0.24) | 2.00(0.23) | 0.912 |
| Ever tobacco smoker, n(%) | 1,097(29.8) | 908(29.6) | 189(30.9) | 0.509 |
| Hypertension, n(%) | 2,703(73.5) | 2,242(73.1) | 461(75.5) | 0.225 |
| Systolic blood pressure(SD), mmHg | 127.6(18.7) | 127.6(18.6) | 127.9(19.0) | 0.735 |
| Diastolic blood pressure (SD), mmHg | 75.0(10.9) | 75.1(11.0) | 74.6(10.3) | 0.231 |
| Heart rate(SD), bpm | 64.0(10.7) | 64.0(10.7) | 63.8(10.6) | 0.676 |
| QRS duration(SD), ms | 89.0(15.6) | 89.0(15.7) | 89.1(15.3) | 0.798 |
| QT interval(SD), ms | 416.5(30.6) | 416.3(31.5) | 417.6(32.9) | 0.396 |
| Bazett corrected QT (SD), ms | 426.7(25.8) | 426.6(25.7) | 427.2(26.3) | 0.608 |
| Framingham corrected QT (SD),ms | 422.2(22.7) | 422.0(22.5) | 422.9(23.5) | 0.423 |
| Hodge corrected QT (SD), ms | 423.5(23.0) | 423.3(22.7) | 424.2(24.4) | 0.408 |
| Fridericia corrected QT (SD), ms | 423.0(22.1) | 422.9(22.6) | 423.7(23.6) | 0.435 |
| Cornell voltage(SD), μV | 1514(597) | 1507(593) | 1547(618) | 0.146 |
| Median beat: normal sinus, n(%) | 3,629(99) | 3,026(98.6) | 603(98.7) | 0.573 |
| Median beat: atrial fibrillation, n(%) | 36(1.0) | 13(0.4) | 1(0.2) |  |
| Median beat: ventricular pacing, n(%) | 14(0.4) | 29(1.0) | 7(1.2) |  |
| QRS area(SD), mV*ms | 38.4(18.3) | 38.4(18.3) | 38.7(18.3) | 0.659 |
| Peak QRS magnitude(SD), mV | 1.59(0.44) | 1.59(0.44) | 1.60(0.45) | 0.395 |
| Area QRS azimuth(95%CI),° | 20.9(20.1-21.6) | 20.7(19.9-21.6) | 21.5(19.5-23.4) | 0.008 |
| Peak QRS azimuth(95%CI),° | 9.7(8.9-10.4) | 9.6(8.8-10.4) | 9.8(8.0-11.7) | >0.05 |
| Area QRS elevation(95%CI),° | 73.3(72.8-73.9) | 73.1(72.5-73.7) | 74.4(72.9-75.8) | >0.05 |
| Peak QRS elevation(95%CI),° | 72.3(71.8-72.7) | 72.2(71.7-72.7) | 72.9(71.8-74.0) | >0.05 |
| T area(SD), mV*ms | 48.5(23.1) | 48.5(22.9) | 48.3(24.0) | 0.835 |
| Peak T magnitude(SD), mV | 0.36(0.16) | 0.36(0.16) | 0.35(0.16) | 0.591 |
| Area T azimuth(95%CI),° | -45.1(-46.1 to -44.1) | -45.2(-46.3 to -44.2) | -44.5(46.8 to -42.2) | >0.05 |
| Peak T azimuth(95%CI),° | -36.2(-37.2 to -35.1) | -36.3(-37.4 to -35.2) | -35.4(-37.9 to -33.0) | >0.05 |
| Area T elevation(95%CI),° | 75.6(75.1-76.1) | 75.6(75.0-76.1) | 76.0(74.8-77.2) | >0.05 |
| Peak T elevation(95%CI),° | 69.9(69.4-70.4) | 69.9(69.4-70.4) | 69.8(68.7-71.0) | >0.05 |
| Wilson's (area) SVG (SD), mV*ms | 69.3(28.2) | 69.4(28.2) | 68.7(28.3) | 0.531 |
| Peak SVG magnitude (SD), mV | 1.81(0.50) | 1.81(0.50) | 1.81(0.50) | 0.797 |
| Area SVG azimuth (95%CI),° | -14.0(-14.7 to -13.3) | -14.1(-14.9 to -13.3) | -13.3(-15.1 to -11.5) | >0.05 |
| Peak SVG azimuth (95%CI),° | 3.7(3.0-4.4) | 3.6(2.9-4.4) | 3.9(2.2-5.7) | >0.05 |
| Area SVG elevation (95%CI),° | 71.5(71.1-72.0) | 71.3(70.8-71.9) | 72.6(71.4-73.9) | >0.05 |
| Peak SVG elevation (95%CI),° | 70.8(70.4-71.2) | 70.7(70.2-71.1) | 71.4(70.3-72.5) | >0.05 |
| SAI QRST(SD), mV*ms | 153.9(51.4) | 153.7(51.2) | 154.9(52.6) | 0.591 |
| VM QT integral (SD), mV*ms | 102.9(34.3) | 102.8(34.1) | 103.5(35.4) | 0.642 |
| Area QRS-T angle(95%CI),° | 67.1(66.0-68.3) | 67.1(65.9-68.3) | 67.2(64.3-70.0) | >0.05 |
| Peak QRS-T angle(95%CI),° | 48.3(47.2-49.4) | 48.4(47.2-49.6) | 48.0(45.2-50.7) | >0.05 |

**Supplemental Table 3.** Beta-coefficients for selected variables in VCG-based prediction models

| Input variable | OLS | Ridge | Elastic net | Lasso | Adaptive | Plug-in |
| --- | --- | --- | --- | --- | --- | --- |
| Age, y | 0.026 | 0.268 | 0.304 | 0.332 | 0.341 | 0.042 |
| Male | 0.072 | 0.026 | - | - | - | - |
| Weight, kg | -0.028 | 0.004 | - | - | - | - |
| Height, cm | 0.097 | -0.002 | - | - | - | - |
| BMI, kg/m<sup>2</sup> | 0.169 | 0.055 | 0.051 | 0.053 | 0.094 | - |
| BMI 3 categories | -0.230 | -0.042 | -0.028 | -0.024 | - | - |
| BSA, m<sup>2</sup> | -3.49 | -0.002 | - | - | - | - |
| Ever tobacco smoker | 0.426 | 0.166 | 0.172 | 0.176 | 0.202 | - |
| Hypertension | 0.756 | 0.239 | 0.258 | 0.289 | 0.331 | - |
| Systolic blood pressure, mmHg | 0.010 | 0.115 | 0.088 | 0.075 | 0.105 | - |
| Diastolic blood pressure, mmHg | -0.024 | -0.176 | -0.151 | -0.138 | -0.177 | - |
| Heart rate, bpm | -0.113 | 0.032 | 0.039 | 0.040 | 0.092 | - |
| QRS duration, ms | 0.003 | 0.087 | 0.088 | 0.091 | 0.096 | - |
| QT interval ms | -0.018 | -0.030 | - | - | - | - |
| Bazett corrected QT, ms | 0.246 | 0.045 | 0.024 | 0.016 | - | - |
| Framingham corrected QT, ms | 0.086 | 0.005 | - | - | - | - |
| Hodge corrected QT, ms | - | -0.015 | - | - | - | - |
| Fridericia corrected QT, ms | -0.316 | 0.021 | - | - | - | - |
| Cornell voltage, μV | -0.0005 | -0.118 | -0.100 | -0.093 | -0.134 | - |
| Median beat type (3 categories) | 0.172 | 0.026 | 0.015 | 0.011 | - | - |
| Mean RR' interval, ms | 0.026 | -0.027 | -0.035 | -0.034 | -0.002 | - |
| QRS area, μV*ms | -0.00002 | -0.029 | - | - | - | - |
| Peak QRS magnitude, μV | 0.0003 | 0.081 | 0.073 | 0.063 | 0.097 | - |
| Area QRS azimuth,° | 0.001 | -0.009 | - | - | - | - |
| Peak QRS azimuth,° | 0.0008 | 0.004 | - | - | - | - |
| Area QRS elevation,° | -0.005 | -0.022 | - | - | - | - |
| Peak QRS elevation,° | 0.015 | 0.096 | 0.102 | 0.087 | 0.124 | - |
| T area, μV*ms | -0.00003 | -0.065 | -0.030 | -0.028 | -0.019 | - |
| Peak T magnitude, μV | 0.0007 | -0.003 | - | - | - | - |
| Area T azimuth,° | 0.002 | 0.113 | 0.125 | 0.131 | 0.162 | - |
| Peak T azimuth,° | 0.0008 | 0.001 | - | - | - | - |
| Area T elevation,° | 0.002 | -0.003 | - | - | - | - |
| Peak T elevation,° | -0.006 | -0.038 | -0.033 | -0.040 | -0.047 | - |
| Wilson's (area) SVG, μV*ms | 0.000001 | 0.010 | - | - | - | - |
| Peak SVG magnitude, μV | -0.0001 | 0.020 | - | - | - | - |
| Area SVG azimuth,° | 0.001 | 0.041 | 0.014 | 0.007 | - | - |
| Peak SVG azimuth,° | -0.002 | -0.012 | - | - | - | - |
| Area SVG elevation,° | -0.008 | -0.078 | -0.056 | -0.045 | -0.072 | - |
| Peak SVG elevation,° | 0.003 | 0.061 | 0.004 | - | - | - |
| SAI QRST, μV*ms | -0.00001 | 0.009 | - | - | - | - |
| VM QT integral, μV*ms | 0.00004 | 0.035 | - | - | - | - |
| Area QRS-T angle,° | 0.004 | 0.140 | 0.083 | 0.020 | - | - |
| Peak QRS-T angle,° | 0.019 | 0.270 | 0.320 | 0.389 | 0.444 | 0.010 |
| Constant | -24.73 | -2.312 | -2.323 | -2.336 | -2.377 | -5.442 |

**Supplemental Table 4.**
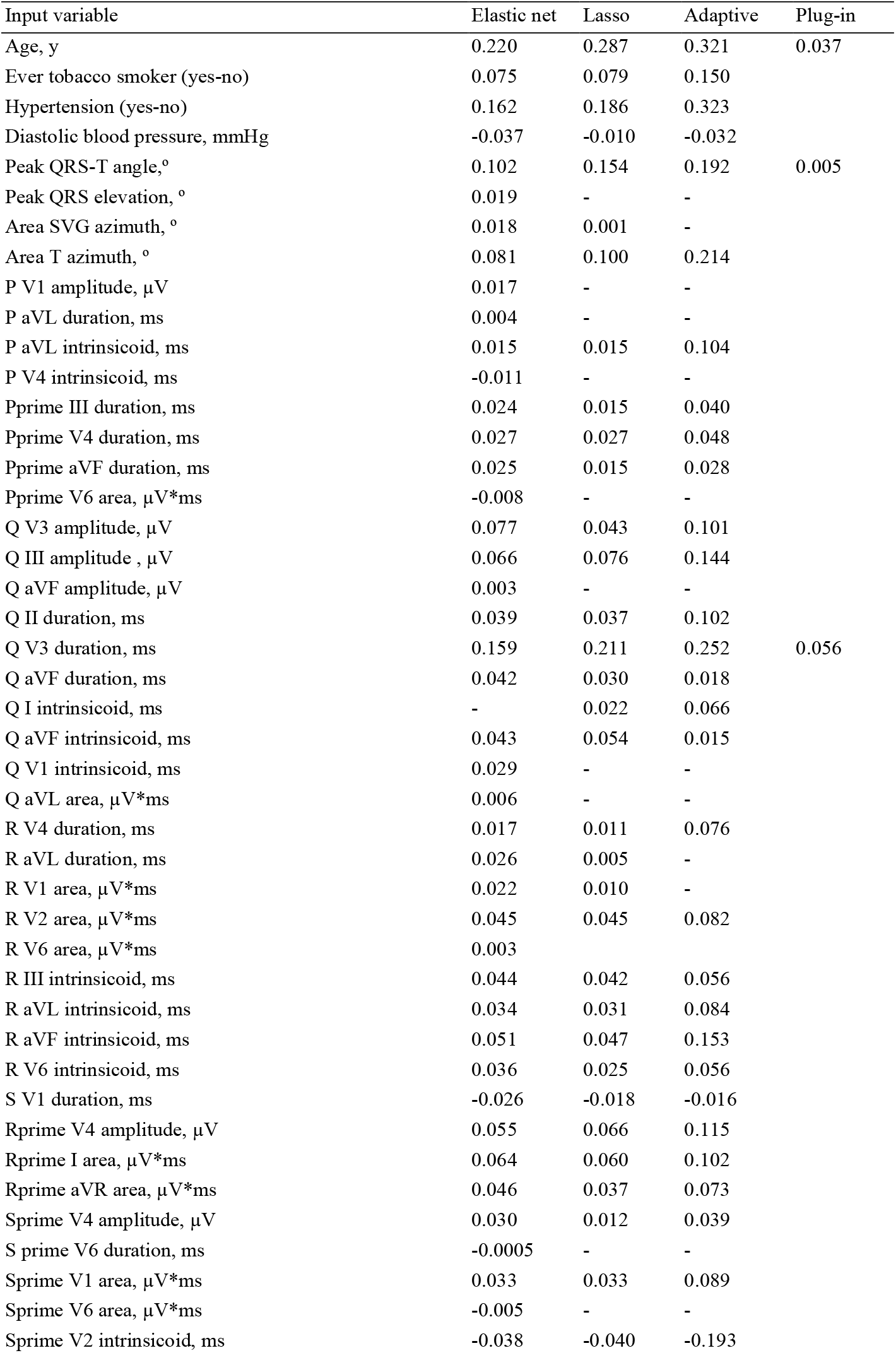
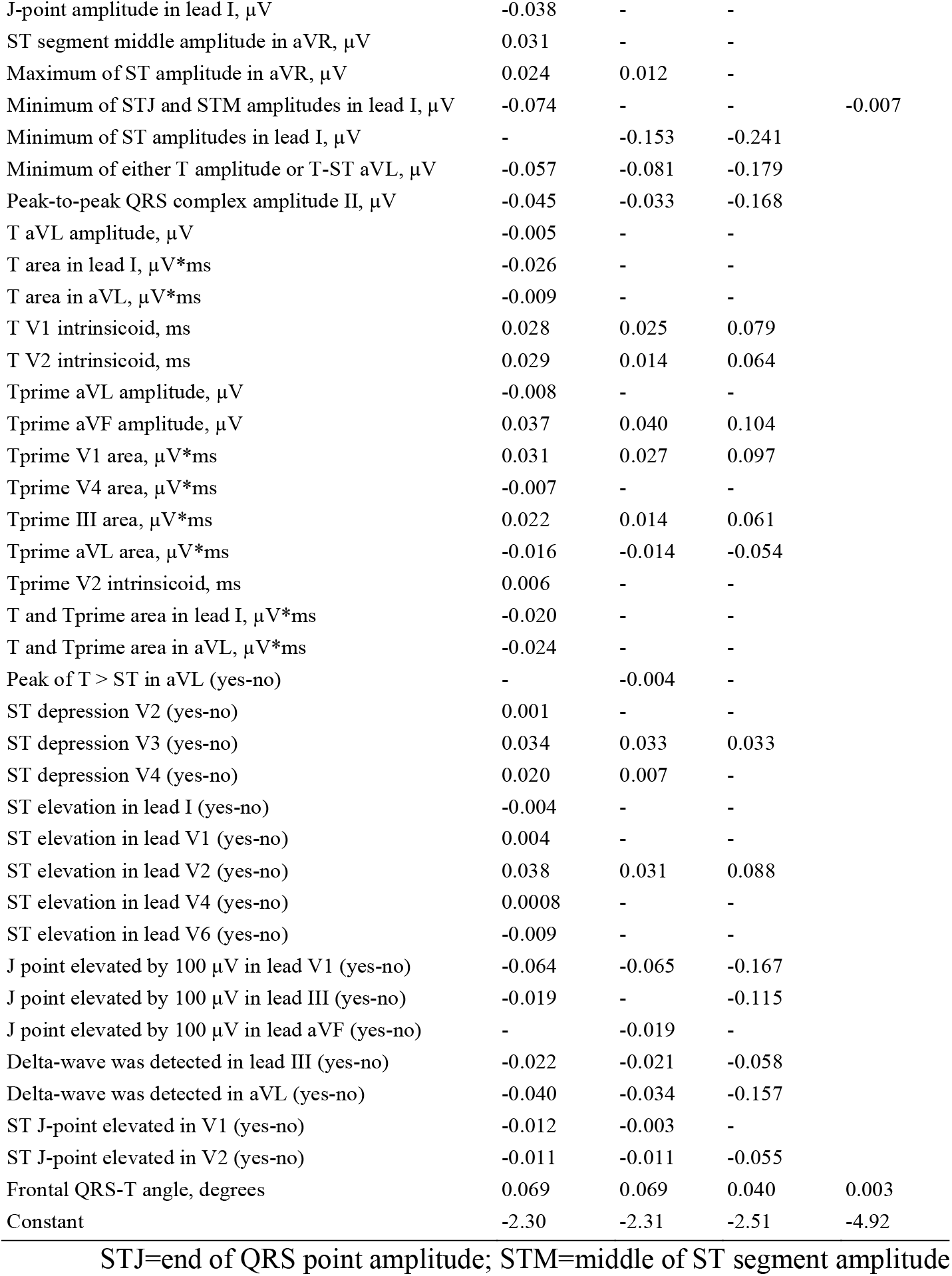
Beta-coefficients for selected variables in VCG+ECG-based models

| Input variable | Elastic net | Lasso | Adaptive | Plug-in |
| --- | --- | --- | --- | --- |
| Age, y | 0.220 | 0.287 | 0.321 | 0.037 |
| Ever tobacco smoker (yes-no) | 0.075 | 0.079 | 0.150 |  |
| Hypertension (yes-no) | 0.162 | 0.186 | 0.323 |  |
| Diastolic blood pressure, mmHg | -0.037 | -0.010 | -0.032 |  |
| Peak QRS-T angle, ° | 0.102 | 0.154 | 0.192 | 0.005 |
| Peak QRS elevation, ° | 0.019 | - | - |  |
| Area SVG azimuth, ° | 0.018 | 0.001 | - |  |
| Area T azimuth, ° | 0.081 | 0.100 | 0.214 |  |
| P V1 amplitude, μV | 0.017 | - | - | |
| P aVL duration, ms | 0.004 | - | - |  |
| P aVL intrinsicoid, ms | 0.015 | 0.015 | 0.104 |  |
| P V4 intrinsicoid, ms | -0.011 | - | - |  |
| Pprime III duration, ms | 0.024 | 0.015 | 0.040 |  |
| Pprime V4 duration, ms | 0.027 | 0.027 | 0.048 |  |
| Pprime aVF duration, ms | 0.025 | 0.015 | 0.028 |  |
| Pprime V6 area, μV*ms | -0.008 | - | - | |
| Q V3 amplitude, μV | 0.077 | 0.043 | 0.101 | |
| Q III amplitude, μV | 0.066 | 0.076 | 0.144 | |
| Q aVF amplitude, μV | 0.003 | - | - | |
| Q II duration, ms | 0.039 | 0.037 | 0.102 |  |
| Q V3 duration, ms | 0.159 | 0.211 | 0.252 | 0.056 |
| Q aVF duration, ms | 0.042 | 0.030 | 0.018 |  |
| Q I intrinsicoid, ms | - | 0.022 | 0.066 |  |
| Q aVF intrinsicoid, ms | 0.043 | 0.054 | 0.015 |  |
| Q V1 intrinsicoid, ms | 0.029 | - | - |  |
| Q aVL area, μV*ms | 0.006 | - | - | |
| R V4 duration, ms | 0.017 | 0.011 | 0.076 |  |
| R aVL duration, ms | 0.026 | 0.005 | - |  |
| R V1 area, μV*ms | 0.022 | 0.010 | - | |
| R V2 area, μV*ms | 0.045 | 0.045 | 0.082 | |
| R V6 area, μV*ms | 0.003 | | | |
| R III intrinsicoid, ms | 0.044 | 0.042 | 0.056 |  |
| R aVL intrinsicoid, ms | 0.034 | 0.031 | 0.084 |  |
| R aVF intrinsicoid, ms | 0.051 | 0.047 | 0.153 |  |
| R V6 intrinsicoid, ms | 0.036 | 0.025 | 0.056 |  |
| S V1 duration, ms | -0.026 | -0.018 | -0.016 |  |
| Rprime V4 amplitude, μV | 0.055 | 0.066 | 0.115 | |
| Rprime I area, μV*ms | 0.064 | 0.060 | 0.102 | |
| Rprime aVR area, μV*ms | 0.046 | 0.037 | 0.073 | |
| Sprime V4 amplitude, μV | 0.030 | 0.012 | 0.039 | |
| S prime V6 duration, ms | -0.0005 | - | - |  |
| Sprime V1 area, μV*ms | 0.033 | 0.033 | 0.089 | |
| Sprime V6 area, μV*ms | -0.005 | - | - | |
| Sprime V2 intrinsicoid, ms | -0.038 | -0.040 | -0.193 |  |
| J-point amplitude in lead I, $\mu\text{V}$ | -0.038 | - | - | |
| ST segment middle amplitude in aVR, $\mu\text{V}$ | 0.031 | - | - | |
| Maximum of ST amplitude in aVR, $\mu\text{V}$ | 0.024 | 0.012 | - | |
| Minimum of STJ and STM amplitudes in lead I, $\mu\text{V}$ | -0.074 | - | - | -0.007 |
| Minimum of ST amplitudes in lead I, $\mu\text{V}$ | - | -0.153 | -0.241 | |
| Minimum of either T amplitude or T-ST aVL, $\mu\text{V}$ | -0.057 | -0.081 | -0.179 | |
| Peak-to-peak QRS complex amplitude II, $\mu\text{V}$ | -0.045 | -0.033 | -0.168 | |
| T aVL amplitude, $\mu\text{V}$ | -0.005 | - | - | |
| T area in lead I, $\mu\text{V}*\text{ms}$ | -0.026 | - | - | |
| T area in aVL, $\mu\text{V}*\text{ms}$ | -0.009 | - | - | |
| T V1 intrinsicoid, ms | 0.028 | 0.025 | 0.079 |  |
| T V2 intrinsicoid, ms | 0.029 | 0.014 | 0.064 |  |
| Tprime aVL amplitude, $\mu\text{V}$ | -0.008 | - | - | |
| Tprime aVF amplitude, $\mu\text{V}$ | 0.037 | 0.040 | 0.104 | |
| Tprime V1 area, $\mu\text{V}*\text{ms}$ | 0.031 | 0.027 | 0.097 | |
| Tprime V4 area, $\mu\text{V}*\text{ms}$ | -0.007 | - | - | |
| Tprime III area, $\mu\text{V}*\text{ms}$ | 0.022 | 0.014 | 0.061 | |
| Tprime aVL area, $\mu\text{V}*\text{ms}$ | -0.016 | -0.014 | -0.054 | |
| Tprime V2 intrinsicoid, ms | 0.006 | - | - |  |
| T and Tprime area in lead I, $\mu\text{V}*\text{ms}$ | -0.020 | - | - | |
| T and Tprime area in aVL, $\mu\text{V}*\text{ms}$ | -0.024 | - | - | |
| Peak of T > ST in aVL (yes-no) | - | -0.004 | - |  |
| ST depression V2 (yes-no) | 0.001 | - | - |  |
| ST depression V3 (yes-no) | 0.034 | 0.033 | 0.033 |  |
| ST depression V4 (yes-no) | 0.020 | 0.007 | - |  |
| ST elevation in lead I (yes-no) | -0.004 | - | - |  |
| ST elevation in lead V1 (yes-no) | 0.004 | - | - |  |
| ST elevation in lead V2 (yes-no) | 0.038 | 0.031 | 0.088 |  |
| ST elevation in lead V4 (yes-no) | 0.0008 | - | - |  |
| ST elevation in lead V6 (yes-no) | -0.009 | - | - |  |
| J point elevated by 100 $\mu\text{V}$ in lead V1 (yes-no) | -0.064 | -0.065 | -0.167 | |
| J point elevated by 100 $\mu\text{V}$ in lead III (yes-no) | -0.019 | - | -0.115 | |
| J point elevated by 100 $\mu\text{V}$ in lead aVF (yes-no) | - | -0.019 | - | |
| Delta-wave was detected in lead III (yes-no) | -0.022 | -0.021 | -0.058 |  |
| Delta-wave was detected in aVL (yes-no) | -0.040 | -0.034 | -0.157 |  |
| ST J-point elevated in V1 (yes-no) | -0.012 | -0.003 | - |  |
| ST J-point elevated in V2 (yes-no) | -0.011 | -0.011 | -0.055 |  |
| Frontal QRS-T angle, degrees | 0.069 | 0.069 | 0.040 | 0.003 |
| Constant | -2.30 | -2.31 | -2.51 | -4.92 |

STJ=end of QRS point amplitude; STM=middle of ST segment amplitude

